# The IDentif.AI 2.0 Pandemic Readiness Platform: Rapid Prioritization of Optimized COVID-19 Combination Therapy Regimens

**DOI:** 10.1101/2021.06.23.21259321

**Authors:** Agata Blasiak, Anh T.L. Truong, Alexandria Remus, Lissa Hooi, Shirley Gek Kheng Seah, Peter Wang, De Hoe Chye, Angeline Pei Chiew Lim, Kim Tien Ng, Swee Teng Teo, Yee-Joo Tan, David Michael Allen, Louis Yi Ann Chai, Wee Joo Chng, Raymond T.P. Lin, David C.B. Lye, John Eu-Li Wong, Gek-Yen Gladys Tan, Conrad En Zuo Chan, Edward Kai-Hua Chow, Dean Ho

## Abstract

**Objectives:** We aimed to harness IDentif.AI 2.0, a clinically actionable AI platform to rapidly pinpoint and prioritize optimal combination therapy regimens against COVID-19.

**Methods:** A pool of starting candidate therapies was developed in collaboration with a community of infectious disease clinicians and included EIDD-1931 (metabolite of EIDD-2801), baricitinib, ebselen, selinexor, masitinib, nafamostat mesylate, telaprevir (VX-950), SN-38 (metabolite of irinotecan), imatinib mesylate, remdesivir, lopinavir, and ritonavir. Following the initial drug pool assessment, a focused, 6-drug pool was interrogated at 3 dosing levels per drug representing nearly 10,000 possible combination regimens. IDentif.AI 2.0 paired prospective, experimental validation of multi-drug efficacy on a SARS-CoV-2 live virus (propagated, original strain, B.1.351 and B.1.617.2 variants) and Vero E6 assay with a quadratic optimization workflow.

**Results:** Within 3 weeks, IDentif.AI 2.0 realized a list of combination regimens, ranked by efficacy, for clinical go/no-go regimen recommendations. IDentif.AI 2.0 revealed EIDD-1931 to be a strong candidate upon which multiple drug combinations can be derived.

**Conclusions:** IDentif.AI 2.0 rapidly revealed promising drug combinations for clinical translation. It pinpointed dose-dependent drug synergy behavior to play a role in trial design and realizing positive treatment outcomes. IDentif.AI 2.0 represents an actionable path towards rapidly optimizing combination therapy following pandemic emergence.

**Graphical Abstract:** 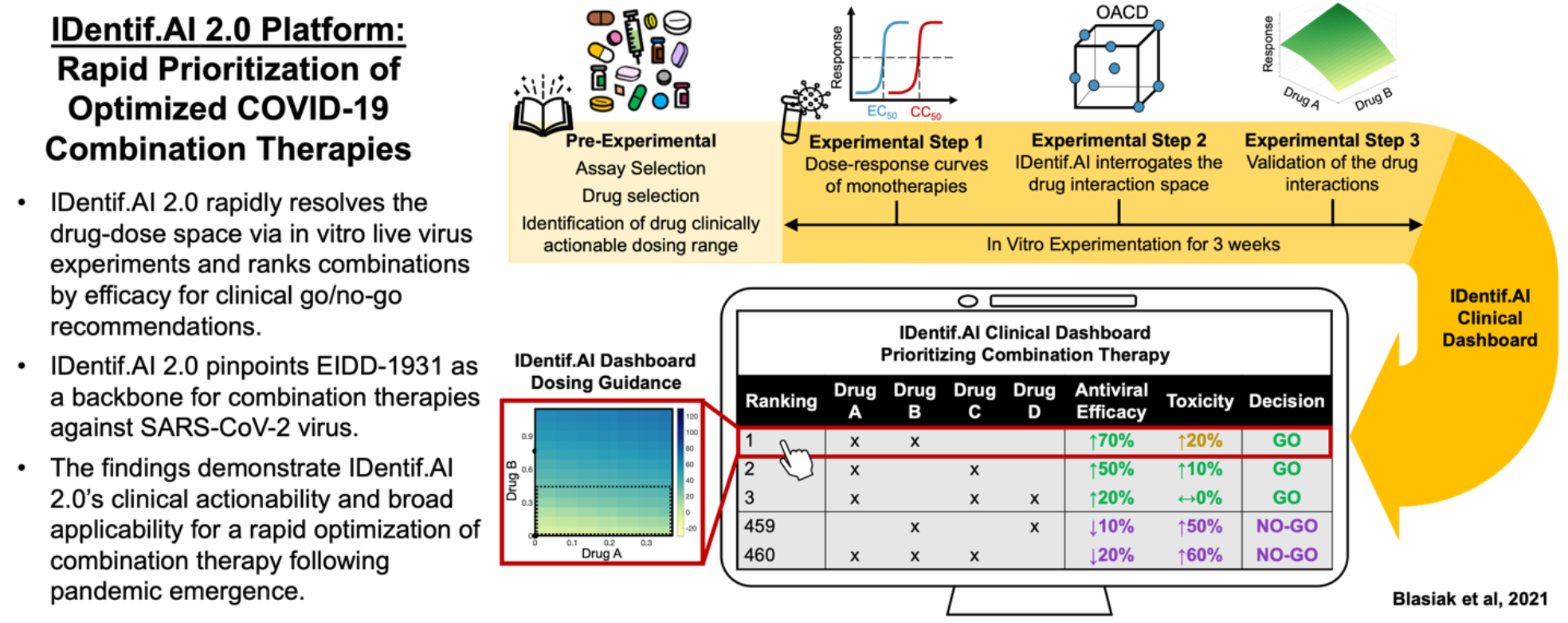

**Highlights:**

- When novel pathogens emerge, the immediate strategy is to repurpose drugs.
- Good drugs delivered together in suboptimal combinations and doses can yield low or no efficacy, leading to misperception that the drugs are ineffective.
- IDentif.AI 2.0 does not use in silico modeling or pre-existing data.
- IDentif.AI 2.0 pairs optimization with prospectively acquired experimental data using a SARS-CoV-2/Vero E6 assay.
- IDentif.AI 2.0 pinpoints EIDD-1931 as a foundation for optimized anti-SARS-CoV-2 combination therapies.

## Introduction

COVID-19 drug development has largely focused on repurposing, either through single agent or combination therapy (Group, 2021, Group et al., 2021; Kalil et al., 2021; Mirabelli et al., 2020; Riva et al., 2020). Clinical trial outcomes of the repurposed candidates were varied (Sheahan et al., 2020; Tekin et al., 2018; White et al., 2021). While many monotherapies did not mediate substantial clinical benefit, their use in properly designed drug combinations may lead to unforeseen efficacy. Addressing this point is challenging for traditional antiviral susceptibility assays. Therefore, developing new methods that leverage unpredictable drug interactions to resolve the complexity of drug selection and dose-dependent drug synergy is essential. In fact, drug and dose selection are so tightly connected that among a pool of candidate therapies, true optimization often yields combinations of unforeseen but clinically acceptable drugs and doses.

Unfortunately, simultaneous drug and dose optimization represents an insurmountable challenge. 12 drugs assessed at 3 dosage levels results in over 500,000 possible combinations. Important strategies for synergy prediction and higher-order drug interaction analysis have been explored (Galindez et al., 2021; Mohapatra et al., 2020; Mongia et al., 2021; Tekin et al., 2018; Zimmer et al., 2016). To address the challenge of ensuring clinical actionability of the combination design outcome, we developed the IDentif.AI platform, an AI-based workflow for rapid combination therapy development. The first permutation of IDentif.AI used neural networks to reveal that the biological response to therapy can be represented by a smooth surface. Subsequent studies resolved this surface, which can rapidly identify optimal combinations, using a second-order algebraic function, with its coefficients determined through a small number of prospective experiments (Abdulla et al., 2020; Al-Shyoukh et al., 2011; Blasiak et al., 2021; Clemens et al., 2019; Ho, 2020; Ho et al., 2020; Lee et al., 2017; Lim et al., 2020; Mohd Abdul Rashid et al., 2015; Rashid et al., 2018; Silva et al., 2016; Wang et al., 2015; Wong et al., 2008). This correlation has subsequently been verified in prospective, human studies in infectious disease, cancer therapy, transplant medicine, and other indications (Tan BKJ et al., 2021; Blasiak et al., 2020; de Mel et al., 2020; Kee et al., 2019; Pantuck et al., 2018; Shen et al., 2020; Zarrinpar et al., 2016). IDentif.AI does not use pre-existing data for algorithm training, in silico modeling, or synergy prediction. Instead, it uses experimental assays to determine the drugs and doses that constitute globally optimized combination regimens. Our previous IDentif.AI studies pinpointed top-ranked combinations (based on inhibition efficacy) mediated by unforeseen drug interactions (Blasiak et al., 2021). Following this study, we developed IDentif.AI 2.0 to expand the resolution of drug and dose analysis to yield a broader spectrum of clinically actionable combinations.

Due to the strong dependence of drug selection and drug dosing on combination therapy optimization, IDentif.AI and IDentif.AI 2.0 utilized clinically relevant dosing levels as reference points for the design of their respective studies. Furthermore, their optimization capabilities have been foundational towards their strong alignment with clinical trial outcomes. In this study (Fig. 1), a starting pool of candidate therapies that have been or are being evaluated in various COVID-19 clinical settings was developed in consultation with the clinical community (Table 1, (Beigel et al., 2020; Goldman et al., 2020; Richardson et al., 2020). IDentif.AI 2.0 implementation on a propagated, original live SARS-CoV-2 strain was completed within 3 weeks. This workflow rapidly pinpointed EIDD-1931 in combination with remdesivir (RDV), EIDD-1931 in combination with baricitinib (BRT), and EIDD-1931 in combination with masitinib (MST) as promising regimens for further development. EIDD-1931/RDV resulted in the highest efficacy and broad synergy across varying dosing levels. EIDD-1931 in combination with BRT or MST showed a stronger dose-dependence on drug synergy compared to EIDD/RDV. These results were reconfirmed against the SARS-CoV-2 B.1.351 (Beta) variant and B.1.617.2 (Delta) variant. The results demonstrate the need for dose optimization studies should these regimens be advanced to clinical trials. The resulting highly ranked combinations also demonstrate that EIDD-1931 (and likely the EIDD-2801 prodrug) is a strong candidate upon which multiple drug combinations can be derived (Cox et al., 2020; Sheahan et al., 2020).

**Table 1.**
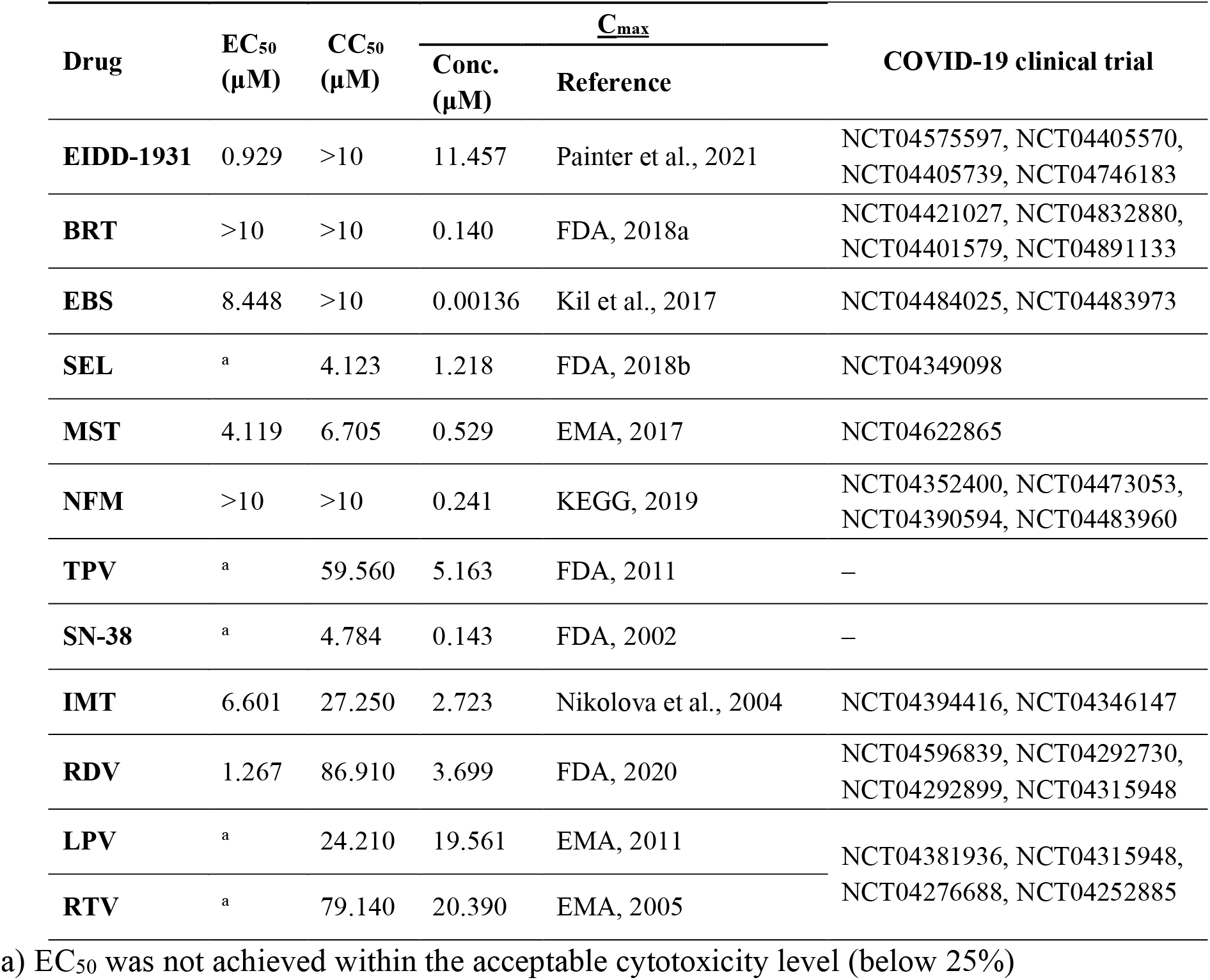
Drug anti-SARS-CoV-2 efficacy and cytotoxicity when administered in monotherapy as compared to C_max_ obtained from the literature and regulatory documents. Absolute EC_50_ and CC_50_ were obtained from the D-R curves for each drug individually constructed based on a CPE viral assay with Vero E6 cells. EC curves were plotted after excluding %Inhibition values corresponding to drug concentrations resulting in %Cytotoxicity above 25%. Baricitinib (BRT), ebselen (EBS), selinexor (SEL), masitinib (MST), nafamostat mesylate (NFM), telaprevir (VX-950) (TPV), imatinib mesylate (IMT), remdesivir (RDV), lopinavir (LPV), and ritonavir (RTV).

**Fig. 1.**
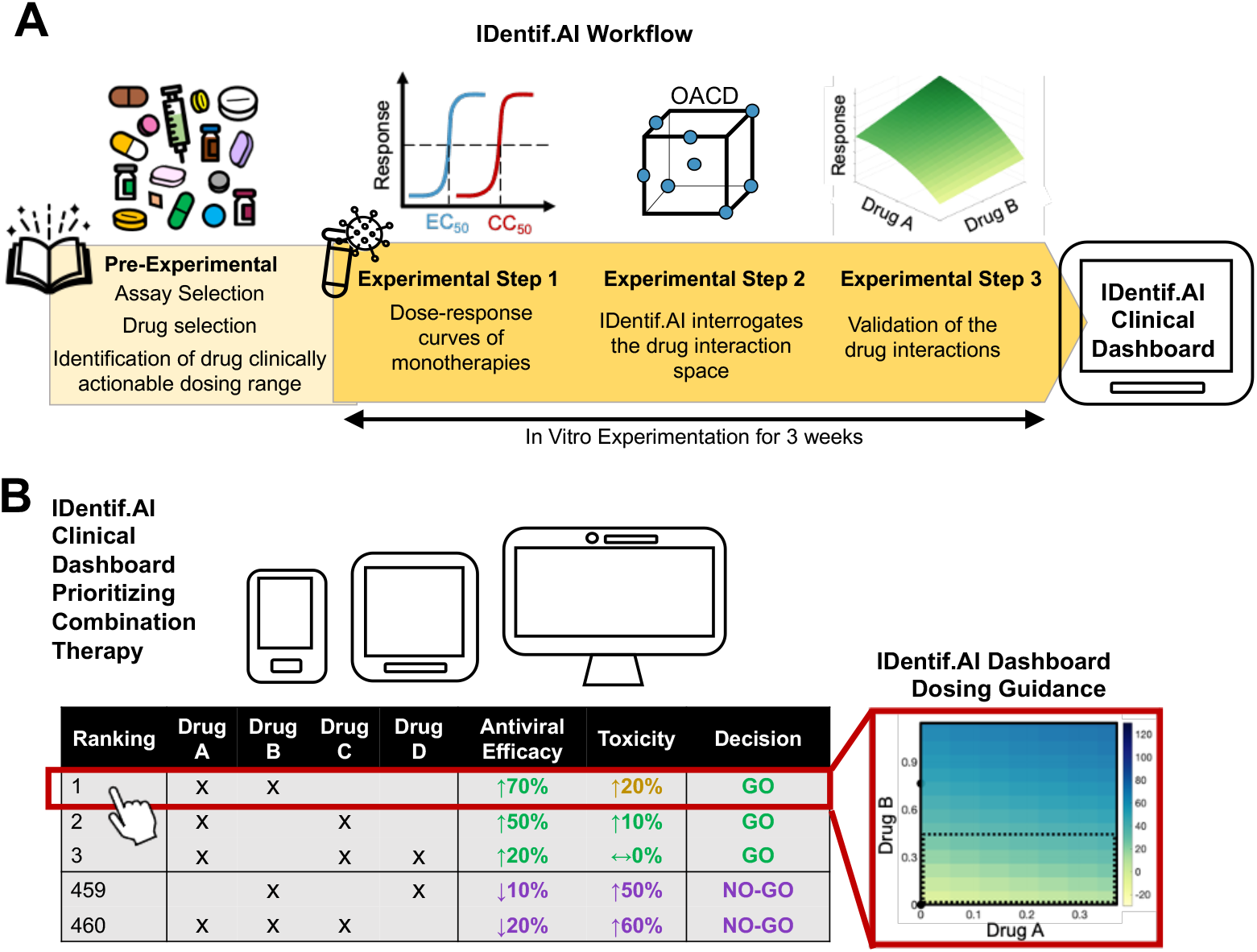
IDentif.AI workflow and actionable outcome. A) IDentif.AI workflow for optimizing drug combinations. Clinical applicability considerations are integrated into IDentif.AI workflow to pre-emptively best position the optimized combinations for a clinical translation. B) The outcome of the IDentif.AI workflow is a list of ranked combinations for prioritizing drug combinations for go/no-go decisions.

IDentif.AI 2.0 did not rely on pre-existing datasets, synergy predictions, or in silico modeling to design these combinations. Instead, it harnessed experimental data from carefully designed drug-dose permutations and prospectively executed studies to drive the optimization process to complement existing strategies in the fight against COVID-19 pandemic. These results demonstrate the potential actionability of IDentif.AI 2.0 as an effective go/no-go platform for prioritizing and advancing combination therapies towards further preclinical or clinical development, and clinical decision support system (CDSS).

## Materials and Methods

Additional sections can be found in Supplemental Materials and Methods.

### Viral inhibition and cell cytotoxicity of drugs

All experiments with the live SARS-CoV-2 (propagated, original strain, B.1.351 and B.1.617.2 variants) were performed in a BSL-3 laboratory. Each treatment was prepared in the culture media and pipetted into the wells of the white 96-well plate in triplicate. 2 × 10^4^ Vero E6 cells were added into each well with and without SARS-CoV-2 (100 TCID_50_) to test viral CPE inhibition and cytotoxicity effects, respectively. The maximum DMSO concentration used in each experimental step and media only served as vehicle controls. Plates were incubated for 72 h before measuring the cell viability via Viral ToxGlo (Promega, G8941) per manufacturer’s instructions. Drug cytotoxicity and viral CPE inhibition were calculated, as described previously (Blasiak et al., 2021). In case no difference was detected between the vehicle and cell only controls, the results from these treatments in each plate were pooled together and served as plate-specific control used in the calculations. The calculations used in the validation experimental step used an average of pooled measurements from the control treatments from all plates. GraphPad Prism 9 software (GraphPad Software) was used to plot dose-response (D-R) curves and to derive absolute effective concentrations EC_10_, EC_20_ and EC_50_ of %Inhibition and absolute cytotoxic concentrations CC_50_ of %Cytotoxicity.

%Cytotoxicity in the validation experimental step was calculated in THLE-2 human liver and AC16 human cardiomyocyte cell lines. The drugs were added to the wells after the cells were allowed to adhere to the surface for 24 h. The plates were incubated for 72 h before measuring the cell viability via CellTiter-GLO (Promega, G7570) per manufacturer’s instructions. %Cytotoxicity calculations in THLE-2 and AC16 were performed using an average of pooled measurements from the control treatments from all plates.

### Drug interaction analysis in the IDentif.AI experimental step

IDentif.AI analysis correlated the 6-drug in vitro experimental data into a quadratic series to elicit optimized drug combinations and drug-drug interactions. The analysis was performed in MATLAB R2020a (Mathworks, Inc.) (Blasiak et al., 2021). IDentif.AI analysis derived two quadratic series – %Inhibition, %Cytotoxicity – by including all experimental replicates as inputs and performing bidirectional elimination in which the *P* value from the F-statistic served as the removal criterion. Box-Cox transformation determined appropriate transformations to improve the residual distributions and the goodness of the fit represented by adjusted R^2^. Residual analysis was performed for each of the two IDentif.AI-derived series. Outlier analysis was performed based on the residual distribution.

### Drug interaction analyses in the validation experimental step

Interaction surfaces were constructed using drug combinations selected via D-optimal experimental design (N = 9 treatments for EIDD-1931/RDV and EIDD-1931/BRT; N = 6 treatments for EIDD-1931/MST; 3-4 replicates per treatment) performed in MATLAB R2020a (MathWorks, Inc). We assumed a quadratic model of the drug interactions. All replicates were included in the construction of the surfaces.

GraphPad Prism 9 software (GraphPad Software) was used to plot D-R curves and derive EC_50_ of %Inhibition and CC_50_ of %Cytotoxicity of the validation set treatments (monotherapies and combinations). Drug combinations were tested at two fixed ratios: i) Level 2/Level 2 ratio for EIDD-1931/RDV, and Level 1/Level 2 ratio for EIDD-1931/BRT from IDentif.AI experimental set (OACD ratio); and ii) C_max_/C_max_ ratio (C_max_ ratio). %Cytotoxicity was evaluated in terms of its effects on the %Inhibition assay.

### Statistical analyses

All in vitro experiments were performed in at least 3 biological replicates. %Inhibition and %Cytotoxicity are presented as mean ± propagated standard deviation (SD) (Supplementary Materials and Methods). The IDentif.AI-estimated coefficients were analyzed using sum of squares F-test and *P*-values for each individual coefficient obtained from stepwise regression.

## Results

### Starting drug pool and live virus/in vitro experimental model

The 12 candidate drugs had hypothesized mechanisms of either inhibiting SARS-CoV-2 entry into the host cell– BRT, NFM, IMT– or inhibiting SARS-CoV-2 replication – EIDD-1931, EBS, SEL, MST, TPV, SN-38, RDV, LPV, RTV (Drayman et al., 2020; El Bairi et al., 2020; Haritha et al., 2020; Kneller et al., 2020; Lovetrue, 2020; Sanders et al., 2020; Sheahan et al., 2020; Stebbing et al., 2020; Xiu et al., 2020).

The antiviral drug effects in monotherapies and in combinations were tested by exposing Vero E6 cells for 72 h to SARS-CoV-2 before measuring the %Inhibition of the virus-induced cytopathic effect (CPE) and the drug toxicity-induced CPE (%Cytotoxicity). The %Inhibition and %Cytotoxicity were derived in independent biological replicates based on different activity ranges, so their effect sizes are not directly comparable. The Z’-factor of 0.569 (N = 52 positive controls and 70 negative controls) indicated that across all three experimental steps (without experimental sets retesting the results in SARS-CoV-2 variants), the separation between the negative and positive controls was sufficient to perform an ‘excellent’ assay (Zhang et al., 1999). Assay quality details for each experimental step are included in the Supplementary Results.

### Monotherapies were broadly not sufficiently efficacious in the actionable dosing range

The first experimental step aimed to gauge the drugs’ antiviral activities when the drugs were administered as monotherapies. We exposed the Vero E6 cells with and without the live virus to an increasing concentration of each drug on its own, constructed D-R curves and calculated half maximal absolute effective concentration (EC_50_) – the drug concentration at which half of the viral-induced CPE is inhibited. An analogical process was performed to understand at what concentration each drug became cytotoxic.

Importantly, to ensure the clinical actionability of the findings, the concentration range tested for each drug was selected with consideration of their maximum plasma concentration (C_max_) achieved in the human body (Table 1) to capture the efficacy in a concentration range of interest - clinically actionable concentrations with potential human efficacy. A high C_max_/EC_50_ ratio indicates a drug’s capability to reach the concentrations in the human blood plasma that is sufficient to provide antiviral efficacy (Arshad et al., 2020). The specifics of the C_max_ selection for each drug are presented in the Supplementary Results.

The D-R curves (Fig. S1) revealed that the antiviral activities of the drugs were limited when they were administered as monotherapies (Table 1). RDV and EIDD-1931 were the only drugs that achieved EC_50_ < C_max_ with C_max_ /EC_50_ ratios of 2.92 and 11.46, respectively. RDV, RTV and LPV performance in monotherapy was comparable with that observed in the previous IDentif.AI study based on the same assay (Blasiak et al., 2021).

### IDentif.AI 2.0 drug combination optimization

IDentif.AI was developed as a CDSS for real-world application under pandemic preparedness circumstances, where experimentation is often performed under shortened timelines. In addition, IDentif.AI can be executed in concert with high biosafety level laboratories and specified viral volumes processed per session. While the IDentif.AI 2.0 workflow substantially reduces the time and workload needed for combination design compared to traditional methods, natural biological and experimental variations resulted in an additional study team oversight process in the workflow of narrowing the drug pool based on promising interaction profiles observed from the initial 12-drug experiment. This additional step selected RDV, EBS, MST, IMT, BRT and EIDD-1931 to be included in the focused, 6-drug IDentif.AI experimental set that enabled the team to complete the downstream optimization process alongside laboratory guidelines while also minimizing biological and experimental variation. As IDentif.AI 2.0 aims to serve as a CDSS in a real-life setting, its potential for scalable and widespread deployment is a key consideration.

In the focused, 6-drug IDentif.AI experimental step, Vero E6 cells with and without the live virus were exposed to the drug treatments in monotherapies and according to a 50-combination Orthogonal Array Composite Design (OACD) table (Table S1). The drugs in the treatments had 3 concentration levels (Table 2). 10% C_max_ for each drug was broadly considered as an achievable dose at the target tissue and served as the maximum concentration level. EIDD-1931 concentration was further restricted to EC_20_ to avoid overrepresentation of this drug in the experimental set and a potential saturation of the %Inhibition results (Table 2).

**Table 2.**
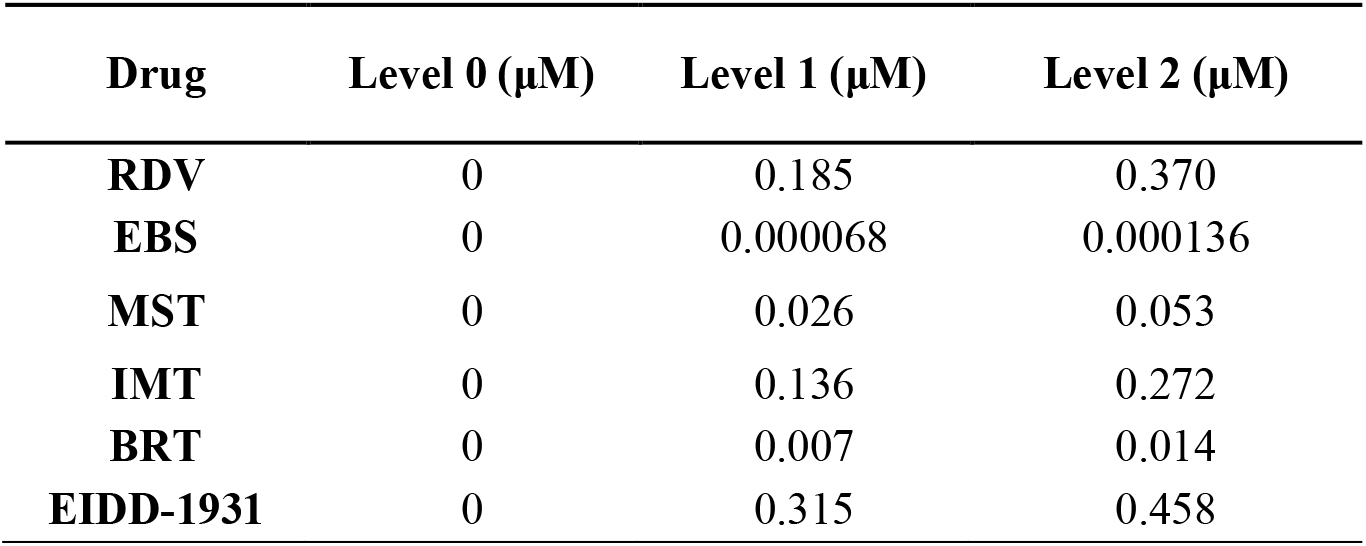
Clinically actionable drug concentrations for the IDentif.AI 2.0 drug combination optimization. Concentration Level 0 indicated lack of the drug, concentration Level 1 and Level 2 were selected based on 5% and 10% C_max_ for RDV, EBS, MST, IMT, BRT. Concentration Level 1 and Level 2 were selected based on absolute EC_10_ and absolute EC_20_ for EIDD-1931. Remdesivir (RDV), ebselen (EBS), masitinib (MST), imatinib mesylate (IMT), baricitinib (BRT).

| **Drug** | **Level 0 ($\mu\text{M}$)** | **Level 1 ($\mu\text{M}$)** | **Level 2 ($\mu\text{M}$)** |
| --- | --- | --- | --- |
| **RDV** | 0 | 0.185 | 0.370 |
| **EBS** | 0 | 0.000068 | 0.000136 |
| **MST** | 0 | 0.026 | 0.053 |
| **IMT** | 0 | 0.136 | 0.272 |
| **BRT** | 0 | 0.007 | 0.014 |
| **EIDD-1931** | 0 | 0.315 | 0.458 |

IDentif.AI 2.0 analysis used a quadratic equation to describe the 6-drug interaction space against the SARS-CoV-2 (adjusted R^2^ = 0.794; Table S2). Monotherapy results demonstrated that EIDD-1931 was the most efficacious drug in the pool, even when given at EC_20_, with moderate antiviral effects. IDentif.AI 2.0 analysis of the drug-drug interaction detected an unforeseen interaction between EIDD-1931 and RDV, which was the most efficacious two-drug combination, and were predicted to achieve close to maximal %Inhibition in a synergistic interaction demonstrated by the convex shape of the EIDD-1931/RDV interaction surface (Fig. 2). In addition, IDentif.AI-derived coefficients pointed to an interaction between EIDD-1931 and BRT. The EIDD-1931/BRT interaction surface had a mildly concave shape across the tested BRT concentration range. BRT was predicted to have a mild antagonistic effect on the EIDD-1931-driven %Inhibition at its mid-concentration (Fig. 2). Little to no cytotoxic effects have been detected in the IDentif.AI experimental step (Supplementary Results).

**Fig. 2.**
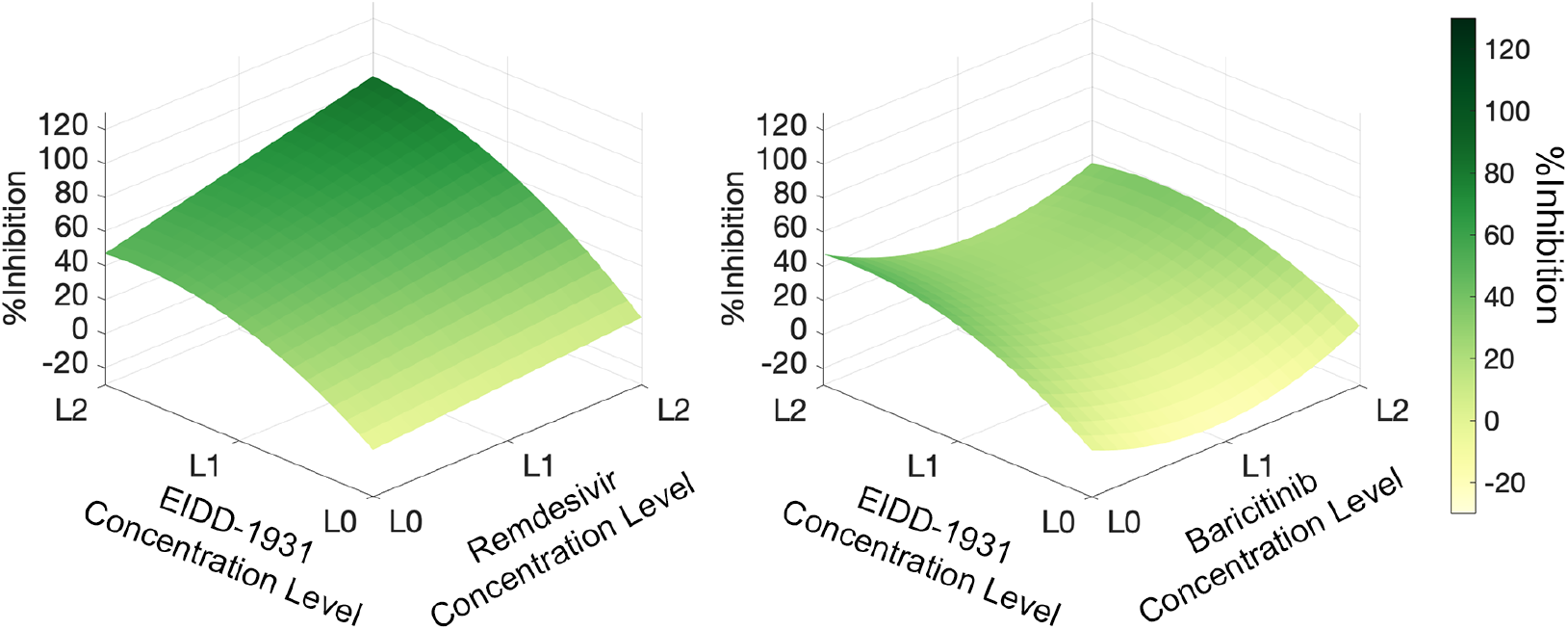
IDentif.AI interaction analysis. The analysis indicates that EIDD-1931 interacts differently with remdesivir and baricitinib. EIDD-1931/remdesivir interaction surface had a convex shape indicating a synergistic interaction, while EIDD-1931/baricitinib had a concave interaction surface had a concave shape indicating a dose-dependent, mildly antagonistic interaction. L0, L1, L2 correspond to concentration Level 0, Level 1 and Level 2.

### Experimental validation of the IDentif.AI analysis

In the IDentif.AI 2.0 validation step we exposed Vero E6 cells to live virus to construct interaction surfaces for EIDD-1931/RDV and EIDD-1931/BRT assuming a quadratic equation. With the focus on a two-drug interaction only, we recalibrated the size of the validated interaction space to range from 0% to 15% C_max_ of EIDD-1931 to capture the clinically actionable range (<10% C_max_) and the adjacent space (Fig. 3).

**Fig. 3.**
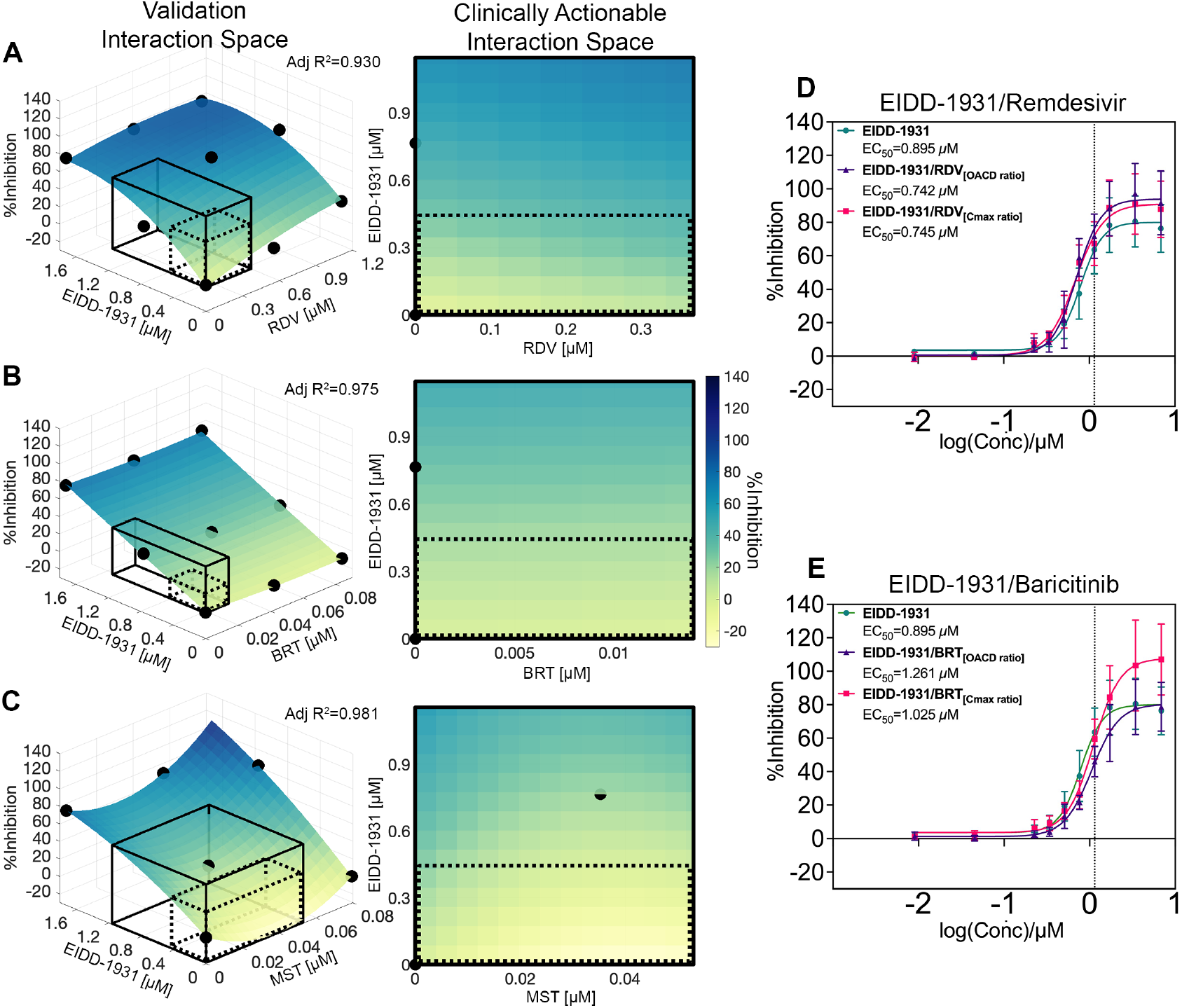
Validation of EIDD-1931 drug interactions affecting %Inhibition in the propagated, original SARS-CoV-2 strain. (A-C) Surface plots of EIDD-1931 interactions with remdesivir (RDV), baricitinib (BRT) and masitinib (MST) in the validation interaction space, clinically actionable interaction space (black, solid line border) and the interaction space from the IDentif.AI 2.0 analysis (black, dotted line border). The latter two are also shown as 2-dimenisional maps. All experiments were performed with N = 3 to 4 replicates, which were independently included in the surface construction. Black, round markers indicate an average %Inhibition of the replicates for each treatment. Adjusted R^2^ (Adj R^2^) indicates goodness of the fit for each interaction surface. (D-E) Dose-response curves (D-R curves) of EIDD-1931 in monotherapy and in a combination with RDV and BRT at two concentration ratios: the ratio tested in the IDentif.AI 2.0 experimental set (OACD ratio) and the ratio dictated by the C_max_ values of the drugs (C_max_ ratio). Half maximal absolute effective concentration (EC_50_) was derived from the D-R curves. The vertical line marks the 10% C_max_ of EIDD-1931. Please note that the EIDD-1931-only EC_50_ values (Green) were provided in both subfigures D and E to enable direct comparisons with both combinations (EIDD-1931/RDV and EIDD-1931/BRT). The entire assay was completed in 1 experiment, realizing all data points in a single global study and enabling comprehensive derivation of combinations and direct comparisons between monotherapies and combinations. Error bars represent propagated SD (N = 3 to 4 replicates). Of note, this propagated SD did not arise from the replicates’ spread, but from plate-to-plate variation (SD of the controls).24

The EIDD-1931/RDV interaction surface had a convex shape pointing to the highest %Inhibition achieved when both drugs are at their highest concentrations, suggesting it is beneficial to provide these drugs in a combination. The flat shape of the EIDD-1931/BRT interaction surface indicated that the %Inhibition results driven by EIDD-1931 are not affected by the presence of BRT. Due to interesting multi-drug behavior observed from IDentif.AI 2.0 analysis, we further assessed the EIDD-1931/MST combination. Of note, IDentif.AI 2.0 analysis did not identify a significant interaction between EIDD-1931 and MST. After expanding the concentration range in the validation set to 15%C_max_ of both EIDD-1931 and MST, the concave shape of the EIDD-1931/MST interaction surface indicated that an increase in the concentrations of both drugs could mediate maximum %Inhibition. This phenomenon, however, had the strongest effect outside of the clinically actionable range, potentially explaining why the EIDD-1931/MST interaction was not detected in the IDentif.AI 2.0 step.

Given the previously demonstrated immunomodulatory activity of BRT and synergistic potential of MST, the EIDD-1931/BRT and EIDD-1931/MST combinations can potentially be evaluated further, where dose optimization studies may be essential to achieving optimal efficacy.

### Dose-response curves revealed additional information for the EIDD-1931 interactions with RDV and BRT

Interaction surfaces were constructed with a small number of drug combination data points and therefore had a limited resolution. To validate IDentif.AI-determined EIDD-1931 interactions with RDV and BRT at a higher fidelity, in the same data set, we included drug treatments to generate D-R curves at the two different drug ratios: as used in the OACD table and dictated by C_max_.

The D-R curves revealed additional information. We observed a slight shift in the D-R curves for both combinations: towards a lower and higher EIDD-1931’s absolute EC_50_ for EIDD-1931/RDV and EIDD-1931/BRT, respectively (Figs. 3D and E; Fig. S2). The mild antagonistic effect of BRT at the OACD ratio is consistent with the IDentif.AI 2.0 analysis. Overall, these results suggest small effect sizes of the tested interactions of EIDD-1931. Interestingly, at high concentrations, the D-R curve shapes revealed a potential boost in maximum %Inhibition achievable by EIDD-1931 co-administered with RDV or BRT (Fig. 3D and E; Fig. S3). This phenomenon was not observed for EIDD-1931/BRT_[OACD ratio]_ which instead was shown to induce a mildly antagonistic shift in EC_50_. The observation that the same drug combination at two different ratios can potentially exhibit the opposite interactions points out to the importance of optimizing drug doses at the same time as their combinations. Further experiments are required to confirm and better characterize these observations.

The potentially synergistic efficacy interaction demonstrates that combining EIDD-1931 with either RDV or BRT can achieve a higher efficacy than each drug alone, which highlights the potential of EIDD-1931 serving as a backbone to combinational therapies against the SARS-CoV-2. However, as the synergy was detected outside of the actionable interaction space, additional dosing strategies may need to be considered to optimize these interactions in a clinical setting.

### The efficacy of the pinpointed therapies against SARS-CoV-2 B.1.351 and B.1.617.2 variants

We retested the efficacy of the pinpointed monotherapies and combination treatments against the SARS-CoV-2 B.1.351 and B.1.617.2 variants. When tested against B.1.351 variant EIDD-1931 and RDV monotherapies demonstrated an increased antiviral activity as compared to the propagated, original strain (Fig. 4 and S4). Accordingly, the EIDD-1931 interaction surfaces demonstrated saturation regions at high concentrations of EIDD-1931 and RDV (Fig. 4A). When tested against B.1.617.2 variant, EIDD-1931 retained its high antiviral activity, while RDV demonstrated an increased antiviral activity as compared to the propagated, original strain (Fig. 4 and S4). Similar to the propagated, original strain, the effects of EIDD-1931 combinations depended on the ratio in which the drugs were combined. Overall, the experiments with SARS-CoV-2 B.1.351 and B.1.617.2 variants confirm the EIDD-1931 for a consideration as a monotherapy and as a backbone of combinatory treatment against SARS-CoV-2. Dose adjustments in combination therapy should be performed for each specific variant.

**Fig. 4.**
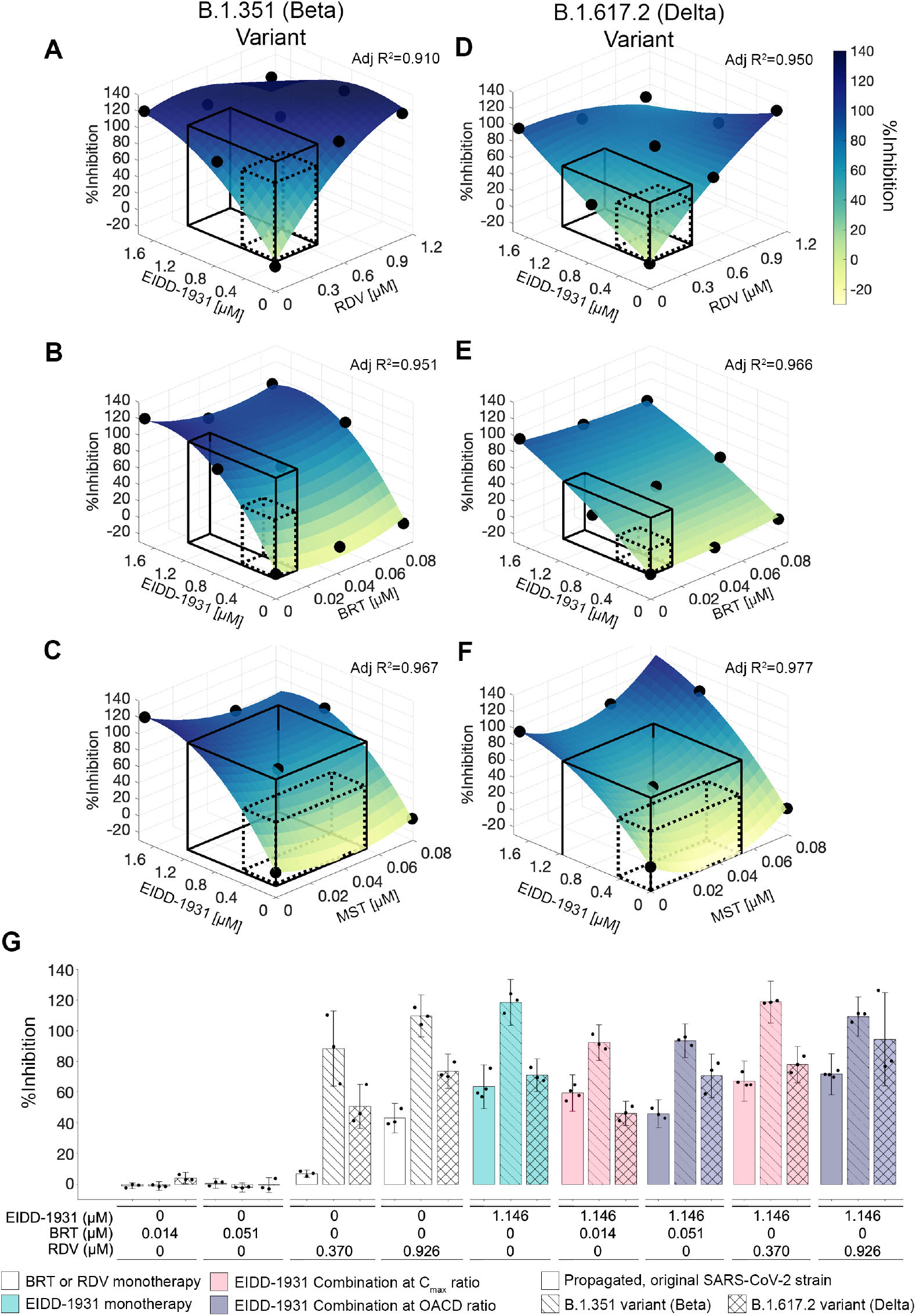
Validation of EIDD-1931 drug interactions affecting %Inhibition in SARS-CoV-2 B.1.351 and B.1.617.2 variants. (A-F) Surface plots of EIDD-1931 interactions with remdesivir (RDV), baricitinib (BRT) and masitinib (MST) in the validation interaction space, clinically actionable interaction space (black, solid line border) and the interaction space from the IDentif.AI 2.0 analysis (black, dotted line border). All experiments were performed with N = 3 replicates, which were independently included in the surface construction. Black, round markers indicate an average %Inhibition of the replicates for each treatment. Adjusted R^2^ (Adj R^2^) indicates goodness of the fit for each interaction surface. The experiments with SARS-CoV-2 B.1.351 variant (A-C), and B.1.617.2 variant (D-F) were performed in two independent sets. (G) %Inhibition against the propagated, original SARS-CoV-2 strain (bars with block filling), B.1.351 variant (bars with line filling) and B.1.617.2 variant (bars with cross lines filling) of 10% C_max_ EIDD-1931 in monotherapy (green) and in a combination with RDV and BRT at two concentration ratios: the ratio dictated by the C_max_ values of the drugs (C_max_ ratio; pink) and the ratio tested in the IDentif.AI 2.0 experimental set (OACD ratio; purple). Black markers indicate individual data points. Error bars represent propagated standard deviation (SD). Of note, this propagated SD did not arise from the replicates’ spread, but from plate-to-plate variation (SD of the controls). No statistically significant difference was detected with Kruskal-Wallis test when followed by Dunn’s post hoc test.

### Cytotoxicity of EIDD-1931 in the interactions

When %Cytotoxicity was tested in VeroE6, the EIDD-1931/RDV and EIDD-1931/BRT interaction surfaces (Fig. 5A and B; Fig. S5) had a convex shape while the EIDD-1931/MST (Fig. 5C) had a concave shape indicating that %Cytotoxicity is a result of an interaction between EIDD-1931 and the drugs. However, %Cytotoxicity was low and was not predicted to expand beyond 23% for any of the drug combinations in the actionable range.

**Fig. 5.**
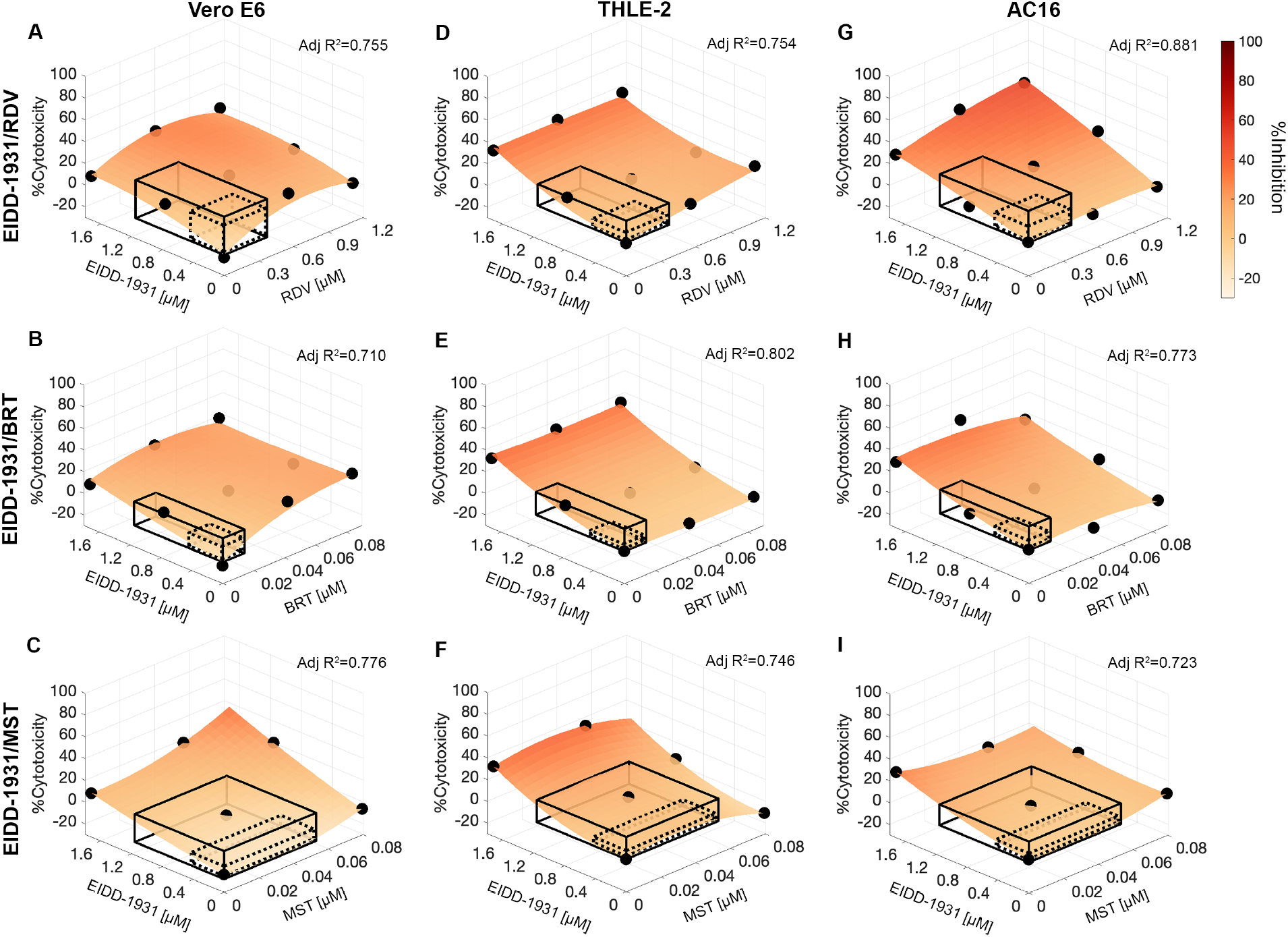
Validation of EIDD-1931 drug interactions affecting %Cytotoxicity. (A-I) Surface plots of EIDD-1931 interactions with remdesivir (RDV), baricitinib (BRT) and masitinib (MST), in the validation interaction space, clinically actionable interaction space (black, solid line border) and the interaction space from the IDentif.AI analysis (black, dotted line border), based on the experimentation in Vero E6 cells (A-C), THLE-2 (D-F) and AC16 (G-I). (A-I) All experiments were performed with N = 3 to 4 replicates, which were independently included in the surface construction. Black, round markers indicate an average %Cytotoxicity of the replicates for each data point. Adjusted R^2^ (Adj R^2^) indicates goodness of the fit for each interaction surface.

To gauge the potential cytotoxicity that may be observed in clinical settings, we investigated cytotoxic effects of the EIDD-1931 drug combinations in cell lines of human origin: liver epithelial cells (THLE-2) and cardiomyocytes (AC16). The interaction surfaces had different shapes in different cell lines, highlighting the target-specific cytotoxic characteristics of the treatments. The regular shape of each interaction surface with uniformly high %Cytotoxicity tested at high EIDD-1931 concentration independent of the presence of the other drugs indicate that cytotoxicity in THLE-2 was driven by EIDD-1931 and it was not significantly affected by its interactions with RDV, BRT and MST (Fig. 5D-F; Fig S5). In AC16 cells, MST did not increase EIDD-1931-driven cytotoxicity; BRT mildly alleviated it in a ratio-dependent fashion; and RDV demonstrated dose-dependent cytotoxic synergy, with predicted 29% maximum %Cytotoxicity in the clinically actionable interaction space (Fig. 5G-I; Fig S5).

## Discussion

### IDentif.AI 2.0 actionability

This work has demonstrated IDentif.AI 2.0’s potential as a clinically actionable platform to rapidly prioritize drug combinations for further consideration based on high efficacy, and de-prioritize combinations that may be avoided due to lack of efficacy as optimized to a specific SARS-CoV-2 variant. A key technical attribute of IDentif.AI 2.0 is that its combination design workflow is inherently based on optimization via drug and dosing data that are clinically relevant.

Our study pinpoints EIDD-1931 to be a promising therapeutic for COVID-19, both as a monotherapy and as the backbone drug for combination therapies. EIDD-1931 is hypothesized to inhibit viral replication by inducing lethal mutagenesis in coronaviruses (Sheahan et al., 2020). EIDD-2801 (EIDD-1931’s prodrug) has been shown to inhibit SARS-CoV-2 in primary human airway epithelial cell cultures and in multiple animal models (Cox et al., 2021; Rosenke et al., 2021; Sheahan et al., 2020; Wahl et al., 2021) and is currently being evaluated in a Phase 3 trial (NCT04575597). Its ability to be administered orally supports its potential as a rapidly deployable therapy (Merck & Co., 2021). Additionally, our toxicity results for EIDD-1931 are in line with other studies that have shown that EIDD-2801 may be well-tolerated (Painter et al., 2021; Khoo SH et al., 2021). The resulting efficacy of the IDentif.AI 2.0 validation combinations indicates that they too may be suitable for further preclinical and/or clinical evaluation with careful dosing adjustment strategy to maximize synergistic efficacy at minimal toxicity.

### Limitations of the study

It is important to note that this study was conducted in an in vitro live virus model, and the subsequent preclinical and clinical dose optimization and evaluation will be needed. This study evaluated a pre-specified drug pool, and further studies with additional drug candidates are warranted. Developing a set of drug selection criteria such as drug class, administration route, prior evidence of interaction with other drugs, clinical relevance and accessibility may streamline the development of the drug pool. While the current study did not examine the immunomodulatory effects of the anti-inflammatories (SN-38, BRT), future work using applicable assays towards combination therapy development with immunomodulators is warranted as IDentif.AI 2.0 can be implemented in virtually all assays, provided quantifiable efficacy and toxicity readouts are available. Including immunomodulation will potentially create viable therapeutic options for severe patients as shown by recent clinical progress (Group et al., 2021).

The IDentif.AI 2.0 workflow has some technical limitations, nonetheless, it is developed for rapid optimization and clinical actionability, and complementary strategies can be integrated to address them. First, the IDentif.AI interaction space interrogation assumes a quadratic relationship with the efficacy/cytotoxicity responses. The optimized combinations presented here are largely limited to two-drug combinations, rapidly identifying the most significant drugs and its partners from a large drug combination search set. Further development of more complex combinatorial therapy strategies, such as four- or five-drug combinations would likely require some reconciliation of higher-order interactions, similar to previous studies (Tekin et al., 2018). Second, only limited dosage ratios were tested. Nevertheless, the current results suggest that further preclinical and clinical dosing optimization may reveal the full potential of the pinpointed combination therapies in term of their synergistic potency (i.e., beneficial dose reduction) and synergistic efficacy interactions (i.e., beneficial increase in maximum efficacy) (Meyer et al., 2019). Additional correlation studies with clinical trial outcomes, when available, will also further determine the applicability of IDentif.AI 2.0 towards go/no-go decisions on combination regimens pinpointed by IDentif.AI 2.0. The IDentif.AI 2.0 process in its current form has resulted in promising outcomes, further development of IDentif.AI 2.0 and its potential integration with other methodologies may further enhance its clinical relevance.

### IDentif.AI 2.0 for clinical decision making

Clinical decision making in response to the COVID-19 pandemic has been dynamically adapting to new information (Martinez-Sanz et al., 2020; Metlay and Armstrong, 2021). With new evidence emerging, Infectious Diseases Society of America (IDSA) COVID-19 treatment guidelines provides updated recommendations for certain monotherapies and combinations depending on severity and setting (Bhimraj A, 2021). Dose optimization has been increasingly recognised as a key therapy optimization element for maximizing public health benefits from the therapeutic solutions of limited supply (Strohbehn et al., 2021). Therefore, it will be important to pinpoint combination regimens that are clinically actionable, both in composition and dosing parameters based on available recommendations. The IDentif.AI 2.0 platform reported here could provide key insights and address gaps regarding how to optimally combine the therapies.

The platform that can rapidly identify treatments for prioritized testing will also be beneficial in the emergence of new viral variants, which potentially affects vaccination and therapeutic efficacies, and also for the treatment of patients that could not be vaccinated, including those who remain ill with evidence of sustained viral replication. Additionally, IDentif.AI can be tailored to generate combinations that address supply chain considerations and local regulations to make the most out of what is available in the geographical and economic feasibility context for a pandemic readiness programme that is inclusive of low-and middle-income countries (LMICs).

## Conclusions

This work reports the application of IDentif.AI 2.0 towards the rapid optimization and prioritization of combination therapy regimens against COVID-19. The IDentif.AI 2.0 optimization process pinpointed EIDD-1931/baricitinib, EIDD-1931/masitinib, and EIDD-1931/remdesivir as regimens that may be suitable for further evaluation and development. IDentif.AI 2.0 did not rely on pre-existing datasets, synergy predictions, or in silico modeling to design these combinations. Instead, it harnessed data from carefully designed drug-dose permutations and prospectively executed studies to drive the optimization process to complement existing strategies in the fight against COVID-19 pandemic. The promising findings from this work support the expansion of IDentif.AI 2.0 towards a broad range of applications in addressing antimicrobial resistance as well as optimized intervention using antiviral, antibiotic, and antifungal therapies.

## Data Availability

Data requests can be addressed to the corresponding authors.

## Author contribution

A.B.: Conceptualization, Methodology, Data Curation, Writing-Original Draft, Writing-Review & Editing, Visualisation, Project Administration. A.T.L.T,: Methodology, Software, Formal Analysis, Data Curation, Writing-Original Draft, Writing-Review & Editing, Visualisation. A.R.: Methodology, Formal Analysis, Data Curation, Writing-Original Draft, Writing-Review & Editing, Visualisation. L.H.: Investigation, Validation, Data Curation, Writing-Review & Editing, Project Administration. S.G.K.S.: Investigation, Validation, Data Curation, Writing-Review & Editing. P.W.: Methodology, Software, Validation, Formal Analysis, Data Curation, Writing - Original Draft, Writing - Review & Editing, Visualization, D.H.C.: Investigation, Validation, Data Curation, Writing-Review & Editing. A.P.C.L.: Investigation, Validation, Data Curation, Writing-Review & Editing. K.T.N.: Investigation, Writing-Review & Editing. S.T.T.: Investigation, Writing-Review & Editing. Y.-J.T.: Investigation, Data Curation, Writing-Review & Editing. D.M.A.: Conceptualization, Writing-Review & Editing. L.Y.A.C.: Conceptualization, Writing-Review & Editing. W.J.C.: Conceptualization, Writing-Review & Editing. R.T.P.L.: Investigation, Validation, Writing-Review & Editing. D.C.B.L.: Conceptualization, Writing-Review & Editing. J.E.-L.W.: Conceptualization, Writing-Review & Editing. G.-Y.G.T.: Methodology, Resources, Supervision, Writing-Review & Editing, Project Administration. C.E.Z.C.: Methodology, Resources, Supervision, Writing-Review & Editing, Project Administration. E.K.-H.C.: Conceptualization, Methodology, Supervision, Writing-Review & Editing, Funding Acquisition. D.H.: Conceptualization, Methodology, Resources, Writing-Original Draft, Writing-Review & Editing, Visualisation, Supervision.

## Data availability statement

The data used in the analysis of this study can be shared on reasonable request to the corresponding author.

## Ethics approval

This work was conducted under study number 2012/00917 approved by the Domain Specific Review Board, as well as approved OSHE/iORC protocol 2020-00494.

## Acknowledgements

The authors gratefully acknowledge the National Public Health Library, National Centre for Infectious Diseases; and Associate Professor Justin Jang Hann Chu.

## Appendix A. Supplementary data

Supplementary material related to this article can be found, in the online version, at doi: XXX

## References

Abdulla A, Wang B, Qian F, Kee T, Blasiak A, Ong YH, et al. Project IDentif.AI: Harnessing Artificial Intelligence to Rapidly Optimize Combination Therapy Development for Infectious Disease Intervention. Adv Therap 2020;10.1002/adtp.202000034:2000034.

Al-Shyoukh I, Yu F, Feng J, Yan K, Dubinett S, Ho CM, et al. Systematic quantitative characterization of cellular responses induced by multiple signals. BMC Syst Biol 2011;5:88.

Arshad U, Pertinez H, Box H, Tatham L, Rajoli RKR, Curley P, et al. Prioritization of Anti-SARS-Cov-2 Drug Repurposing Opportunities Based on Plasma and Target Site Concentrations Derived from their Established Human Pharmacokinetics. Clin Pharmacol Ther 2020;108(4):775–90.

Beigel JH, Tomashek KM, Dodd LE, Mehta AK, Zingman BS, Kalil AC, et al. Remdesivir for the Treatment of Covid-19 - Final Report. N Engl J Med 2020;383(19):1813–26.

Bhimraj A MR, Shumaker AH, Lavergne V, Baden L, Cheng VC, Edwards KM, et.al. Infectious Diseases Society of America Guidelines on the Treatment and Management of Patients with COVID-19. Version 4.3.0. ; 2021. Available from: https://www.idsociety.org/practice-guideline/covid-19-guideline-treatment-and-management/. [Accessed 10th June 2021].

Blasiak A, Khong J, Kee T. CURATE.AI: Optimizing Personalized Medicine with Artificial Intelligence. SLAS Technol 2020;25(2):95–105.

Blasiak A, Lim JJ, Seah SGK, Kee T, Remus A, Chye H, et al. IDentif.AI: Rapidly optimizing combination therapy design against severe Acute Respiratory Syndrome Coronavirus 2 (SARS-Cov-2) with digital drug development. Bioeng Transl Med 2021;6(1):e10196.

Clemens DL, Lee BY, Silva A, Dillon BJ, Maslesa-Galic S, Nava S, et al. Artificial intelligence enabled parabolic response surface platform identifies ultra-rapid near-universal TB drug treatment regimens comprising approved drugs. PLoS One 2019;14(5):e0215607.

Cox RM, Wolf JD, Plemper RK. Therapeutic MK-4482/EIDD-2801 Blocks SARS-CoV-2 Transmission in Ferrets. Res Sq 2020;10.21203/rs.3.rs-89433/v1.

Cox RM, Wolf JD, Plemper RK. Therapeutically administered ribonucleoside analogue MK-4482/EIDD-2801 blocks SARS-CoV-2 transmission in ferrets. Nat Microbiol 2021;6(1):11–8.

de Mel S, Rashid MBM, Zhang XY, Goh J, Lee CT, Poon LM, et al. Application of an ex-vivo drug sensitivity platform towards achieving complete remission in a refractory T-cell lymphoma. Blood Cancer J 2020;10(1):9.

Drayman N, Jones KA, Azizi SA, Froggatt HM, Tan K, Maltseva NI, et al. Drug repurposing screen identifies masitinib as a 3CLpro inhibitor that blocks replication of SARS-CoV-2 in vitro. bioRxiv 2020;10.1101/2020.08.31.274639.

El Bairi K, Trapani D, Petrillo A, Le Page C, Zbakh H, Daniele B, et al. Repurposing anticancer drugs for the management of COVID-19. Eur J Cancer 2020;141:40–61.

EMA 2005. Norvir: European Public Assessment Preport - Scientific Discussion. https://www.ema.europa.eu/en/documents/scientific-discussion/norvir-epar-scientific-discussion_en.pdf

EMA 2011. Kaletra: Summary of Product Characteristics. https://www.ema.europa.eu/en/documents/product-information/kaletra-epar-product-information_en.pdf

EMA 2017. Masipro: Assessment Report. https://www.ema.europa.eu/en/documents/assessment-report/masipro-epar-refusal-public-assessment-report_en.pdf

FDA 2002. Camptosar (irinotecan hydrochloride injection). https://www.accessdata.fda.gov/drugsatfda_docs/label/2002/20571s16lbl.pdf

FDA 2011. INCIVEK. https://www.accessdata.fda.gov/drugsatfda_docs/label/2011/201917lbl.pdf

FDA 2018a. Olumiant: Pharmacology/Toxicology Review 2018a. https://www.accessdata.fda.gov/drugsatfda_docs/nda/2018/207924Orig1s000PharmR.pdf

FDA 2018b.Selinexor: Multi-Discipline Review https://www.accessdata.fda.gov/drugsatfda_docs/nda/2019/212306Orig1s000MultidisciplineR.pdf

FDA 2020. Veklury (remdesdivir) EUA Fact Sheet for Healthcare Providers. https://www.fda.gov/media/137566/download

Galindez G, Matschinske J, Rose TD, Sadegh S, Salgado-Albarrán M, Späth J, et al. Lessons from the COVID-19 pandemic for advancing computational drug repurposing strategies. Nature Computational Science 2021;1(1):33–41.

Goldman JD, Lye DCB, Hui DS, Marks KM, Bruno R, Montejano R, et al. Remdesivir for 5 or 10 Days in Patients with Severe Covid-19. N Engl J Med 2020;383(19):1827–37.

Group PTC. Azithromycin for community treatment of suspected COVID-19 in people at increased risk of an adverse clinical course in the UK (PRINCIPLE): a randomised, controlled, open-label, adaptive platform trial. Lancet 2021;397(10279):1063–74.

Group RC, Horby P, Lim WS, Emberson JR, Mafham M, Bell JL, et al. Dexamethasone in Hospitalized Patients with Covid-19. N Engl J Med 2021;384(8):693–704.

Haritha CV, Sharun K, Jose B. Ebselen, a new candidate therapeutic against SARS-CoV-2. Int J Surg 2020;84:53–6.

Ho D. Artificial intelligence in cancer therapy. Science 2020;367(6481):982–3.

Ho D, Quake SR, McCabe ERB, Chng WJ, Chow EK, Ding X, et al. Enabling Technologies for Personalized and Precision Medicine. Trends Biotechnol 2020;38(5):497–518.

Kalil AC, Patterson TF, Mehta AK, Tomashek KM, Wolfe CR, Ghazaryan V, et al. Baricitinib plus Remdesivir for Hospitalized Adults with Covid-19. N Engl J Med 2021;384(9):795–807.

Kee T, Weiyan C, Blasiak A, Wang P, Chong JK, Chen J, et al. Harnessing CURATE.AI as a Digital Therapeutics Platform by Identifying N-of-1 Learning Trajectory Profiles. Adv Therap 2019;2(9):1900023.

KEGG 2019. Fusan (Nafamostat Mesilate). https://www.kegg.jp/medicus-bin/japic_med?japic_code=00048710

Khoo SH, FitzGerald R, Fletcher T, Ewings S, Jaki T, Lyon R, et al. Optimal dose and safety of molnupiravir in patients with early SARS-CoV-2: a phase 1, dose-escalating, randomised controlled study. medRxiv 2021;10.1101/2021.05.03.21256309.

Kil J, Lobarinas E, Spankovich C, Griffiths SK, Antonelli PJ, Lynch ED, et al. Safety and efficacy of ebselen for the prevention of noise-induced hearing loss: a randomised, double-blind, placebo-controlled, phase 2 trial. Lancet 2017;390(10098):969–79.

Kneller DW, Galanie S, Phillips G, O’Neill HM, Coates L, Kovalevsky A. Malleability of the SARS-CoV-2 3CL M(pro) Active-Site Cavity Facilitates Binding of Clinical Antivirals. Structure 2020;28(12):1313–20 e3.

Lee BY, Clemens DL, Silva A, Dillon BJ, Maslesa-Galic S, Nava S, et al. Drug regimens identified and optimized by output-driven platform markedly reduce tuberculosis treatment time. Nat Commun 2017;8:14183.

Lim JJ, Goh J, Rashid Mbma, Chow EK-H. Maximizing Efficiency of Artificial Intelligence-Driven Drug Combination Optimization through Minimal Resolution Experimental Design. Adv Therap 2020;3(4):1900122.

Lovetrue B. The AI-discovered aetiology of COVID-19 and rationale of the irinotecan+ etoposide combination therapy for critically ill COVID-19 patients. Med Hypotheses 2020;144:110180.

Martinez-Sanz J, Perez-Molina JA, Moreno S, Zamora J, Serrano-Villar S. Understanding clinical decision-making during the COVID-19 pandemic: A cross-sectional worldwide survey. EClinicalMedicine 2020;27:100539.

Merck & Co. I. Merck and Ridgeback Biotherapeutics Provide Update on Progress of Clinical Development Program for Molnupiravir, an Investigational Oral Therapeutic for the Treatment of Mild-to-Moderate COVID-19. Kenilworth, N.J., & Miami, 2021. Available from: https://www.merck.com/news/merck-and-ridgeback-biotherapeutics-provide-update-on-progress-of-clinical-development-program-for-molnupiravir-an-investigational-oral-therapeutic-for-the-treatment-of-mild-to-moderate-covid-19/ [Accessed 10th June 2021].

Metlay JP, Armstrong KA. Clinical Decision Making During the COVID-19 Pandemic. Ann Intern Med 2021;174(5):691–3.

Meyer CT, Wooten DJ, Paudel BB, Bauer J, Hardeman KN, Westover D, et al. Quantifying Drug Combination Synergy along Potency and Efficacy Axes. Cell Syst 2019;8(2):97–108 e16.

Mirabelli C, Wotring JW, Zhang CJ, McCarty SM, Fursmidt R, Frum T, et al. Morphological Cell Profiling of SARS-CoV-2 Infection Identifies Drug Repurposing Candidates for COVID-19. bioRxiv2020;10.1101/2020.05.27.117184.

Mohapatra S, Nath P, Chatterjee M, Das N, Kalita D, Roy P, et al. Repurposing therapeutics for COVID-19: Rapid prediction of commercially available drugs through machine learning and docking. PLoS One 2020;15(11):e0241543.

Mohd Abdul Rashid MB, Toh TB, Silva A, Nurrul Abdullah L, Ho CM, Ho D, et al. Identification and Optimization of Combinatorial Glucose Metabolism Inhibitors in Hepatocellular Carcinomas. J Lab Autom 2015;20(4):423–37.

Mongia A, Saha SK, Chouzenoux E, Majumdar A. A computational approach to aid clinicians in selecting anti-viral drugs for COVID-19 trials. Sci Rep 2021;11(1):9047.

Nikolova Z, Peng B, Hubert M, Sieberling M, Keller U, Ho YY, et al. Bioequivalence, safety, and tolerability of imatinib tablets compared with capsules. Cancer Chemother Pharmacol 2004;53(5):433–8.

Painter WP, Holman W, Bush JA, Almazedi F, Malik H, Eraut N, et al. Human Safety, Tolerability, and Pharmacokinetics of Molnupiravir, a Novel Broad-Spectrum Oral Antiviral Agent with Activity Against SARS-CoV-2. Antimicrob Agents Chemother 2021;10.1128/AAC.02428-20.

Pantuck AJ, Lee D-K, Kee T, Wang P, Lakhotia S, Silverman MH, et al. Modulating BET Bromodomain Inhibitor ZEN-3694 and Enzalutamide Combination Dosing in a Metastatic Prostate Cancer Patient Using CURATE.AI, an Artificial Intelligence Platform. Adv Therap 2018;1(6):1800104.

Rashid M, Toh TB, Hooi L, Silva A, Zhang Y, Tan PF, et al. Optimizing drug combinations against multiple myeloma using a quadratic phenotypic optimization platform (QPOP). Sci Transl Med 2018;10(453).

Richardson P, Griffin I, Tucker C, Smith D, Oechsle O, Phelan A, et al. Baricitinib as potential treatment for 2019-nCoV acute respiratory disease. Lancet 2020;395(10223):e30–e1.

Riva L, Yuan S, Yin X, Martin-Sancho L, Matsunaga N, Pache L, et al. Discovery of SARS-CoV-2 antiviral drugs through large-scale compound repurposing. Nature 2020;586(7827):113–9.

Rosenke K, Hansen F, Schwarz B, Feldmann F, Haddock E, Rosenke R, et al. Orally delivered MK-4482 inhibits SARS-CoV-2 replication in the Syrian hamster model. Nat Commun 2021;12(1):2295.

Sanders JM, Monogue ML, Jodlowski TZ, Cutrell JB. Pharmacologic Treatments for Coronavirus Disease 2019 (COVID-19): A Review. JAMA 2020;323(18):1824–36.

Sheahan TP, Sims AC, Zhou S, Graham RL, Pruijssers AJ, Agostini ML, et al. An orally bioavailable broad-spectrum antiviral inhibits SARS-CoV-2 in human airway epithelial cell cultures and multiple coronaviruses in mice. Sci Transl Med 2020;12(541).

Shen Y, Liu T, Chen J, Li X, Liu L, Shen J, et al. Harnessing Artificial Intelligence to Optimize Long-Term Maintenance Dosing for Antiretroviral-Naive Adults with HIV-1 Infection. Adv Therap 2020;3(4):1900114.

Silva A, Lee BY, Clemens DL, Kee T, Ding X, Ho CM, et al. Output-driven feedback system control platform optimizes combinatorial therapy of tuberculosis using a macrophage cell culture model. Proc Natl Acad Sci U S A 2016;113(15):E2172–9.

Stebbing J, Krishnan V, de Bono S, Ottaviani S, Casalini G, Richardson PJ, et al. Mechanism of baricitinib supports artificial intelligence-predicted testing in COVID-19 patients. EMBO Mol Med 2020;12(8):e12697.

Strohbehn GW, Parker WF, Reid PD, Gellad WF. Socially optimal pandemic drug dosing. Lancet Glob Health 2021; 10.1016/S2214-109X(21)00251-5.

Tan BKJ, Teo CB, Tadeo X, Peng S, Soh HPL, D. SDX, et al. Personalised, Rational, Efficacy-Driven Cancer Drug Dosing via an Artificial Intelligence SystEm (PRECISE): A Protocol for the PRECISE CURATE.AI Pilot Clinical Trial. Front Digit Health 2021;10.3389/fdgth.2021.635524.

Tekin E, White C, Kang TM, Singh N, Cruz-Loya M, Damoiseaux R, et al. Prevalence and patterns of higher-order drug interactions in Escherichia coli. NPJ Syst Biol Appl 2018;4:31.

Wahl A, Gralinski LE, Johnson CE, Yao W, Kovarova M, Dinnon KH, 3rd, et al. SARS-CoV-2 infection is effectively treated and prevented by EIDD-2801. Nature 2021;591(7850):451–7.

Wang H, Lee DK, Chen KY, Chen JY, Zhang K, Silva A, et al. Mechanism-independent optimization of combinatorial nanodiamond and unmodified drug delivery using a phenotypically driven platform technology. ACS Nano 2015;9(3):3332–44.

White KM, Rosales R, Yildiz S, Kehrer T, Miorin L, Moreno E, et al. Plitidepsin has potent preclinical efficacy against SARS-CoV-2 by targeting the host protein eEF1A. Science 2021;371(6532):926–31.

Wong PK, Yu F, Shahangian A, Cheng G, Sun R, Ho CM. Closed-loop control of cellular functions using combinatory drugs guided by a stochastic search algorithm. Proc Natl Acad Sci U S A 2008;105(13):5105–10.

Xiu S, Dick A, Ju H, Mirzaie S, Abdi F, Cocklin S, et al. Inhibitors of SARS-CoV-2 Entry: Current and Future Opportunities. J Med Chem 2020;63(21):12256–74.

Zarrinpar A, Lee DK, Silva A, Datta N, Kee T, Eriksen C, et al. Individualizing liver transplant immunosuppression using a phenotypic personalized medicine platform. Sci Transl Med 2016;8(333):333ra49.

Zhang JH, Chung TD, Oldenburg KR. A Simple Statistical Parameter for Use in Evaluation and Validation of High Throughput Screening Assays. J Biomol Screen 1999;4(2):67–73.

Zimmer A, Katzir I, Dekel E, Mayo AE, Alon U. Prediction of multidimensional drug dose responses based on measurements of drug pairs. Proc Natl Acad Sci U S A 2016;113(37):10442–7.

