## Supplementary Materials for "The IDentif.AI 2.0 Pandemic Readiness Platform: Rapid Prioritization of Optimized COVID-19 Combination Therapy Regimens"

**This PDF file includes:**

Materials and Methods
Equations S1 and S2
Supplementary Results
Figures S1 to S7
Tables S1 to S3
References

1. **Materials and Methods**

#### 1.1. SARS-CoV-2

Biosafety level-3 (BSL-3) laboratory was used to perform all experiments with a live virus. Severe acute respiratory syndrome coronavirus 2 (SARS-CoV-2) isolated from a nasopharyngeal swab in early 2020 under study number 2012/00917 approved by the Domain Specific Review Board (DSRB), as well as approved OSHE/iORC protocol 2020-00494 (Young et al., 2020) was propagated using Vero E6 C1008 cells in minimum Eagle's medium (MEM; Gibco) with 2% heat-inactivated fetal bovine serum (HI-FBS; Gibco). The virus has undergone several rounds of propagation to form the first, SARS-CoV-2 propagated variant used in this study. This propagated variant was identified to have genetic mutations as compared to the original strain. The second viral strain – the B.1.351 (Beta) variant – was isolated from a nasopharyngeal swab in early 2021 in Singapore and was registered in GISAID EpiFlu™ Database under hCoV-19/Singapore/239/2021. The third viral strain – the B.1.617.2. (Delta) variant – was isolated from a nasopharyngeal swab in 2021 in Singapore. Each viral strain was propagated in Vero E6 C1008 cells in MEM with 2% HI-FBS. Viral ToxGlo™ Assay (Promega) was used to determine the virus titre by a standard tissue culture infectious dose (TCID50) endpoint dilution assay and luminescence readout with a microplate reader (BioTek).

#### 1.2. Cell cultures

African green monkey kidney Vero E6 cells (C1008) were cultured in MEM (Gibco) supplemented with 2% HI-FBS prior to the infection and subsequently added in 96-well white plates (Greiner Bio-One) at a density of 2 x $10^4$ cells/well.

The cultivation of the human liver epithelial THLE-2 cells (CRL2706, ATCC) required a coating medium consisting of bronchial epithelial basal medium (BEBM) with human fibronectin (0.01 mg/mL; Biological Industries), bovine collagen Type I (0.03 mg/mL; Stem Cell Technologies) and bovine serum albumin (0.01 mg/mL; Sigma-Aldrich). THLE-2 cells were plated in pre-coated 96-well plates at 3 x $10^3$ cells/well density and cultured in bronchial epithelial cell growth medium (BEGM; Lonza) excluding gentamicin/amphotecirin and epinephrine, but supplemented with 10% FBS (Biowest), human epidermal growth factor (5 ng/mL) and phosphoethanolamine (70 ng/mL).

AC16 human cardiomyocytes (SCC-09, Millipore) were plated in 96-well plates at 2 x $10^3$ cells/well density and cultured in DMEM/F12 (Life Technologies) mixed with 12.5% FBS (Biowest), 1% penicillin-streptomycin (Life Technologies) and L-glutamine (2mm; Life Technologies). All cell cultures were incubated at 37ºC in a humidified atmosphere containing 5% $CO_2$.

#### 1.3. Drugs

EIDD-1931 (Selleck Chemicals, S0833), nafamostat mesylate (NFM; Selleck Chemicals, S1386) and imatinib mesylate (IMT; Selleck Chemicals, S1026) were dissolved in sterile-filtered water. Baricitinib (BRT; Selleck Chemicals, S2851), ebselen (EBS; Selleck Chemicals, S6676), selinexor (SEL; Selleck Chemicals, S7252), masitinib (MST; Selleck Chemicals, S1064), telaprevir (TPV; Selleck Chemicals, S1538), SN-38 (Selleck Chemicals, S4908), remdesivir (RDV; Selleck Chemicals, S8932), lopinavir (LPV; Selleck Chemicals, S1380) and ritonavir (RTV; Selleck Chemicals, S1185) were dissolved in DMSO (MP Biomedicals).

#### 1.4. Drug treatments in each experimental step

The concentration range for each drug in the first experimental step was prepared by a serial dilution with a dilution factor of 3: $1.9 \times 10^{-6}$ μM to 10 μM for EIDD-1931, BRT, EBS, NFM and SN-38; $1.9 \times 10^{-5}$ μM to 100 μM for SEL, MST, TPV, IMT and RDV; $1.1\times 10^{-3}$ μM to 200 μM for LPV and RTV.

In the IDentif.AI experimental step, a set of curated drug combinations consisting of three concentration levels (Level 0, Level 1, and Level 2) for a 6-drug library was designed in accordance with the Orthogonal Array Composite Design (OACD) as described by Xu et al (Xu et al., 2014). Specifically, Level 0 indicated the absence of the drug and Level 1 and Level 2 corresponded to two clinically actionable concentrations of each drug, selected based on the dose-response (D-R) curves and $C_{max}$ values. This 6-drug OACD was generated by combining a resolution VI 32-run two-level fractional factorial and an 18-run three-level orthogonal array. These 50 runs formulated the minimum amount of experimental drug combinations required to screen each drug's effects through their linear, bilinear (drug-drug interactions), and quadratic parameters. The resolution VI 6-drug OACD is tabulated in Table S1.

The treatments in the validation experimental step were performed in 3 to 4 replicates. The concentration ranges for constructing interaction surfaces were set as: 0 - 1.719 μM for EIDD-1931; 0 - 1.10 μM for RDV; 0 - 0.084 μM for BRT and 0 - 0.079 μM for MST to include the concentration ratios explored in the IDentif.AI experimental step at the high EIDD-1931 concentrations.

#### 1.5. Propagated Standard Deviations (SD)

$$\sigma_I^2 = \left(\frac{\partial I}{\partial E_-}\right)^2 \sigma_{E_-}^2 + \left(\frac{\partial I}{\partial c_-}\right)^2 \sigma_c^2 + \left(\frac{\partial I}{\partial c_+}\right)^2 \sigma_{c_+}^2 \quad \textbf{(S1)}$$

$$\sigma_T^2 = \left(\frac{\partial T}{\partial c_+}\right)^2 \sigma_{c_+}^2 + \left(\frac{\partial T}{\partial E_+}\right)^2 \sigma_{E_+}^2 \quad \textbf{(S2)}$$

In equations S1 and S2, $\sigma_T$ and $\sigma_I$ represent the propagated SD for the mean value of %Cytotoxicity and %Inhibition, respectively. The equations consider the spread of the raw luminescence signals of the positive control (control cells), negative control (cells + virus control), and the experimental triplicates with and without virus (cell + drugs + virus and cells + drugs), which are represented by $\sigma_{c+}$, $\sigma_{c-}$, $\sigma_{E+}$, and $\sigma_{E-}$ respectively (Farrance and Frenkel, 2012).

#### 1.6. Statistical Analysis

Sample distribution was tested with Shapiro-Wilk normality test. The Kruskal-Wallis test by ranks was used for multiple comparisons, followed by Dunn's post hoc test for pairwise comparisons.

### 2. Supplementary Results

### 2.1. $C_{max}$ selection

If extensive $C_{max}$ information was available in the literature, the selection was determined by: i) $C_{max}$ resulting from the doses that obtained regulatory approvals, ii) $C_{max}$ at steady state to reflect the drug availability during the multiday SARS-CoV-2 treatment, iii) additionally, if available, we considered the emerging dosages and dosing schedules of the drugs for SARS-CoV-2 treatment as derived from the registered clinical trials. The specifics of the $C_{max}$ selection for each drug is presented below.

Following oral administration of 800 mg bid EIDD-2801 for 5.5 days, its active metabolite EIDD-1931 reached a steady-state $C_{max}$ of 2970 ng/mL (11.457 μM) (Painter et al., 2021). BRT given at 4 mg qd achieved a $C_{max}$ of 52 ng/mL (0.140 μM) (FDA, 2018a). EBS given orally at 600 mg twice daily (bid) for 4 days had a reported $C_{max}$ of 0.372 ng/mL (0.00136 μM) (Kil et al., 2017). The FDA label reported $C_{max}$ of SEL was 540 ng/mL (1.218 μM) following multiple doses of 80 mg on day 1 and 3 of each week for 2 weeks (FDA, 2018b). The steady-state $C_{max}$ of MST was 264 ng/mL (0.529 μM) when administered orally at 200 mg once daily (qd) for 7 days (EMA, 2017). NFM had a reported $C_{max}$ of 130 ng/mL (0.241 μM) when administered intravenously at 0.2 mg/kg/h for 13 to 23 days (KEGG, 2019). The reported $C_{max}$ for TPV was 3510 ng/mL (5.163 μM) after multiple oral doses of 750 mg once every 8 hours with peginterferon alfa and ribavirin in accordance with the FDA label (FDA, 2011). SN-38 had a reported $C_{max}$ of 56 ng/mL (0.143 μM) when a single 340 mg/m$^2$ irinotecan dose was administered intravenously (FDA, 2002). A single oral dose of 400 mg IMT resulted in a $C_{max}$ of 1606 ng/mL (2.723 μM) (Nikolova et al., 2004). According to a recent update by the Food and Drug Administration, RDV achieved a steady-state $C_{max}$ of 2229 ng/mL (3.699 μM) when given intravenously at a 200 mg loading dose followed by 100 mg for 4 or 9 days (FDA, 2020). Notably, this steady-state $C_{max}$ is lower than the $C_{max}$ after a single 200 mg dose (9.0 μM) used in the IDentif.AI study in 2020 (Blasiak et al., 2021). This change reflects the learnings in the one year from the pandemic emergence. LPV was used in combination with RTV at 400/100 mg bid, with a steady-state $C_{max}$ of 12300 ng/mL (19.561 μM) after 2 weeks (EMA, 2011). The reported $C_{max}$ for RTV after a single dose of 600 mg was 14700 ng/mL (20.390 μM) (EMA, 2005).

### 2.2. Assay quality in each experimental step

The quality of the in vitro assay for characterizing the monotherapies was 'excellent' as indicated by Z'- factor of 0.702 (N = 24). No effects of the vehicle on the %Inhibition and %Cytotoxicity were detected when the maximum used vehicle concentration (0.1% DMSO) was compared with the media only cell controls (Student's t-test, N = 24, $P$ = 0.628 and $P$ = 0.092, respectively). We observed that cytotoxicity >25% had effects on the inhibition assay result, therefore we excluded the %Inhibition data points that had >25% corresponding %Cytotoxicity values.

The condensed experimental set had a Z'-factor of 0.590 (N = 12 positive control replicates and N = 18 negative control replicates, respectively), indicating an 'excellent' assay quality. No effect of the vehicle (0.001% DMSO controls) was detected on the %Inhibition and %Cytotoxicity (Wilcoxon rank-sum test, N = 9 vehicle controls and 9 cells in media controls, $P$ = 1 and 1, respectively). Box-Cox transformation was performed on %Inhibition and

%Cytotoxicity data, and a cubic transformation ($T^3$) was applied to improve the residual distributions and the quadratic equations fit, represented by the adjusted $R^2$ value (Fig. S6 and S7). In the performed outlier analysis, we accepted a substantial variation of the data and as such no data point was excluded to represent biological and experimental variation.

The validation experimental set with the propagated SARS-CoV-2 variant had a 'do-able' assay quality as indicated by the Z'-factor of 0.408 (N = 16 positive control replicates and 28 negative control replicates, respectively). No effect of the vehicle (0.006% DMSO controls) was detected on the %Inhibition and %Cytotoxicity in Vero E6 cells (Wilcoxon rank-sum test, N = 15 vehicle controls and 13 cells in media controls, $P = 1$ and 0.711 respectively). None of the %Inhibition data points were excluded based on the %Cytotoxicity as we did not observe the value drop pattern in %Inhibition that would have correlated with %Cytotoxicity indicating the inhibition assay results were not affected by the cytotoxic effects of the drugs. No effect of the vehicle (0.006% DMSO controls) was detected on the %Cytotoxicity in THLE-2 and AC16 cells (Wilcoxon rank-sum test, N = 18, $P = 1$ and 0.142 respective).

The assay quality of the validation experimental sets with the SARS-CoV-2 B.1.351 and B.1.617.2 variants was 'excellent' as indicated by the Z'-factor of 0.576 and 0.503, respectively (N = 8 positive control replicates and 12 negative control replicates for experimental set for each variant). No effect of the vehicle (0.002% DMSO controls) was detected on the %Inhibition (Wilcoxon rank-sum test, N = 8 vehicle controls and 10 cells in media control, $P = 0.302$ and 0.075 respectively). No data points were excluded for the subsequent analyses.

**2.3. %Cytotoxicity in the IDentif.AI experimental step**

IDentif.AI 2.0 analysis also evaluated %Cytotoxicity of the drug combinations to determine their safety. The resulting adjusted $R^2$ value was 0.0173 indicating that a quadratic equation did not have an appropriate fit to describe the %Cytotoxicity in this interaction space (Table S3). This is attributed to an insufficient cytotoxicity effects detected (< 10%Cytotoxicity) when compared to the variation of measurements done in triplicates (average propagated SD = 8.9%) that is inappropriate to perform the analysis with IDentif.AI.

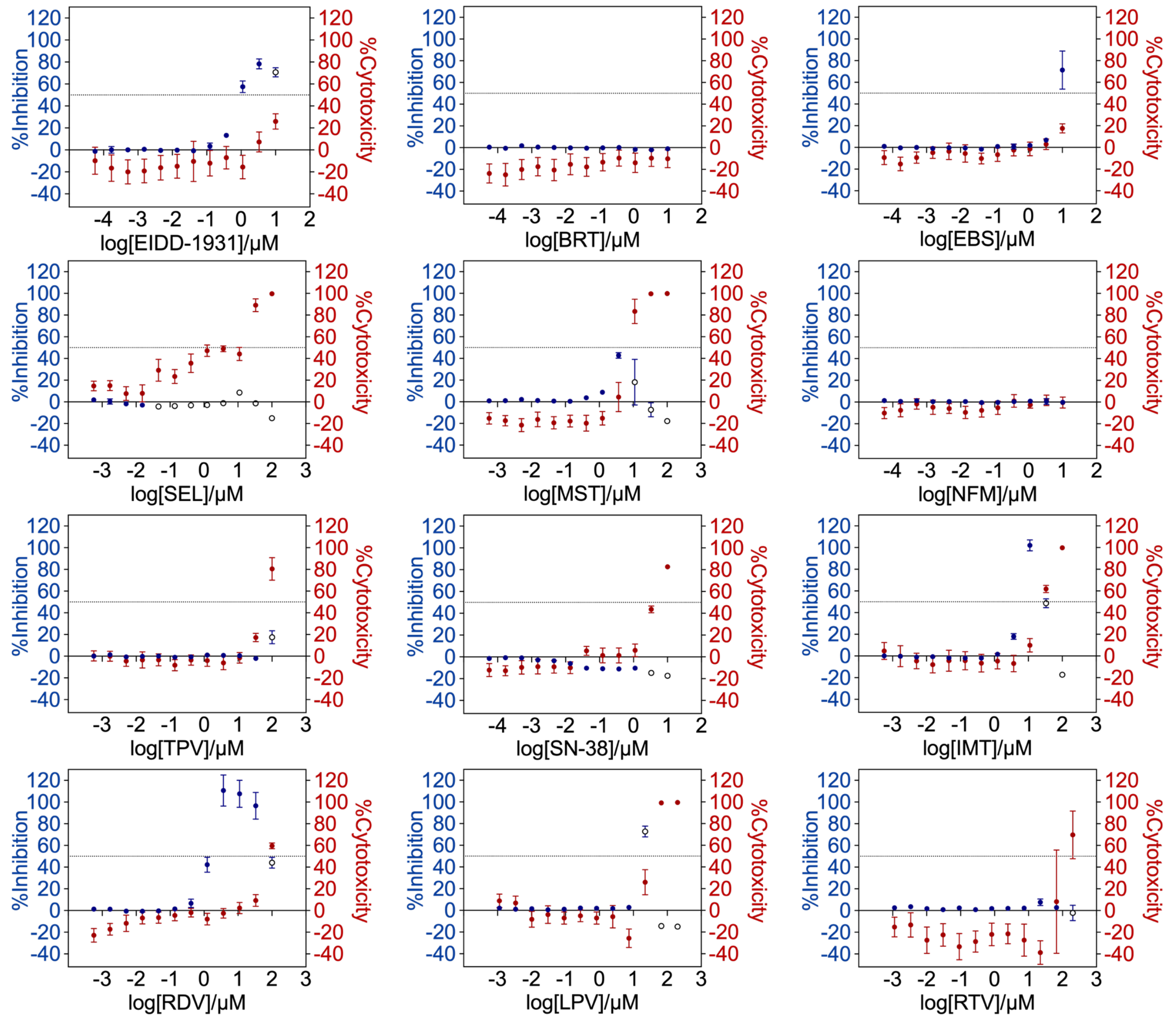


**Fig. S1. Dose-response curves for all 12 selected drugs given as monotherapies.** Vero E6 cells infected with SARS-CoV-2 at 100 $TCID_{50}$ were treated with each drug at different concentrations for 72 h. The viral infection %Inhibition and %Cytotoxicity resulted from these drug treatments in the Vero E6 cells were determined by measuring the luminescence signals of the cell viability in the ATP activity assay. The left and right y-axis represent the %Inhibition (blue) and %Cytotoxicity (red) of the drugs, respectively. The unfilled circles (blue) in the dose-response curves were %Inhibition data points with corresponding toxicities above 25% and were excluded from subsequent dose-response curve analysis. Dotted lines represent absolute $EC_{50}$ and $CC_{50}$, respectively. Data points are mean ± propagated SD (N = 3). Baricitinib (BRT), ebselen (EBS), selinexor (SEL), masitinib (MST), nafamostat mesylate (NFM), telaprevir (VX-950) (TPV), imatinib mesylate (IMT), remdesivir (RDV), lopinavir (LPV), and ritonavir (RTV).

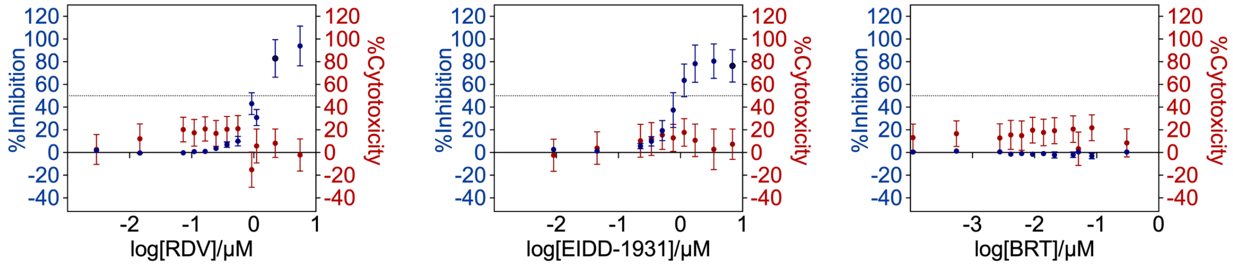


**Fig. S2. Validation dose-response analysis for remdesivir (RDV), EIDD-1931, and baricitinib (BRT).** RDV, EIDD-1931, and BRT at different concentrations were added to Vero E6 cells infected with SARS-CoV-2 at 100 $TCID_{50}$ and incubated for 72 h. The %Inhibition and %Cytotoxicity resulted from the treatments were measured via the luminescence signals of the cell viability in the ATP activity assay. The left and right y-axis of each curve represent the %Inhibition (blue) and %Cytotoxicity (red) for the drugs, respectively. Dotted lines represent absolute $EC_{50}$ and $CC_{50}$, respectively. No data points were excluded in dose-response curve analysis. Data points are presented as mean ± propagated SD (N = 3 to 4).

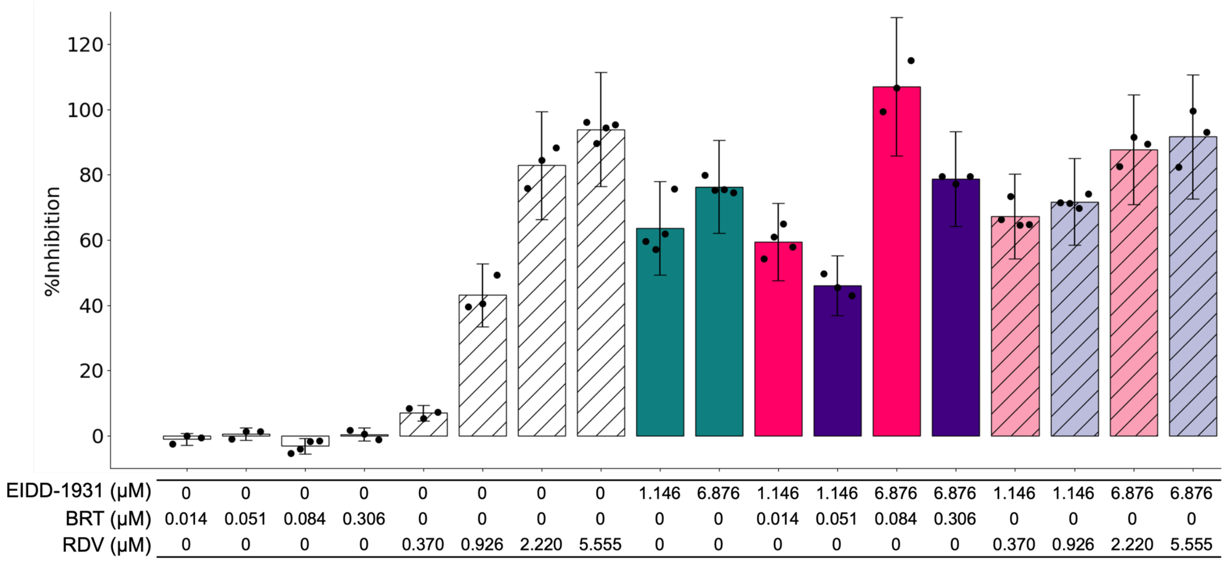


**Fig. S3. Validation of EIDD-1931 interactions at 10% and 60%$C_{max}$ with remdesivir (RDV), and baricitinib (BRT).** EIDD-1931 (green), BRT (white), RDV (white, patterned) and EIDD-1931 combinations at the $C_{max}$ ratio (pink) and at the OACD ratio (purple) at different concentrations were added to Vero E6 cells infected with the propagated, original SARS-CoV-2 strain at 100 $TCID_{50}$ and incubated for 72 h. The %Inhibition resulted from the treatments were measured via the luminescence signals of the cell viability in the ATP activity assay. Data points are presented as mean ± propagated SD (N = 3 to 4). Of note, this propagated SD did not arise from the replicates, but from plate-to-plate variation from control SD. Black, round markers indicate individual replicates. No statistically significant difference was detected with Kruskal-Wallis test when followed by Dunn's post hoc test.

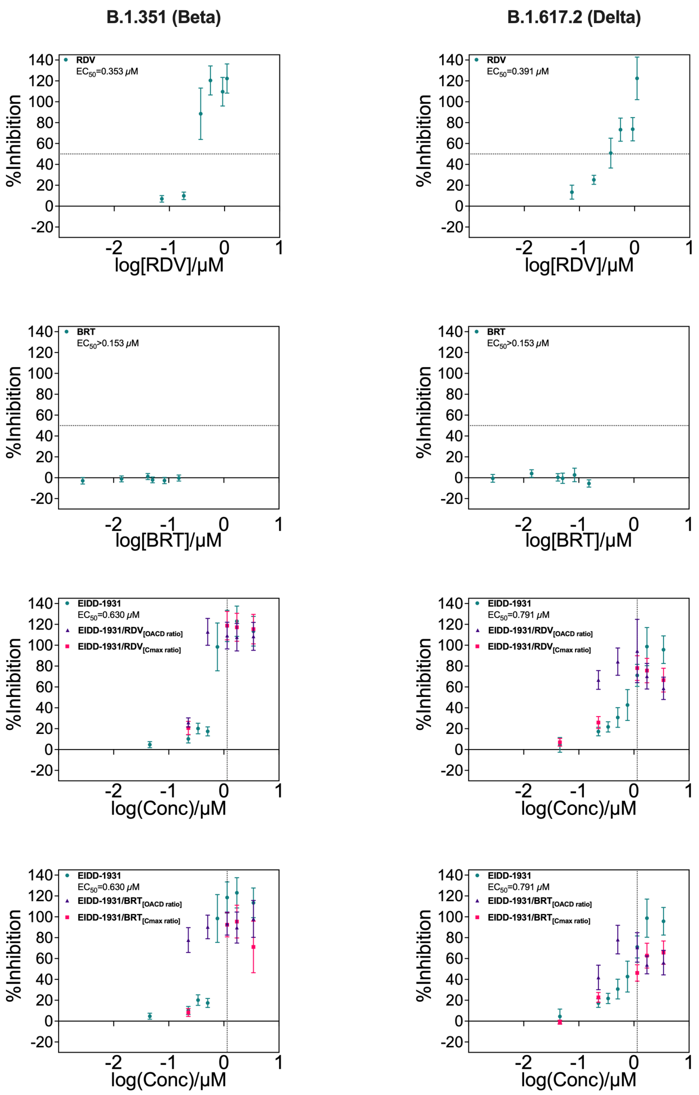


**Fig. S4. Dose-response curves for EIDD-1931, remdesivir (RDV), and baricitinib (BRT) in monotherapies and in combinations against SARS-CoV-2 B.1.351 and B.1.617.2 variants.** Increasing concentrations of EIDD-1931, RDV, and BRT monotherapies were added to Vero E6 cells infected with B.1.351 and B.1.617.2 variants at 100 $TCID_{50}$ and incubated for 72 h. Additionally, selected EIDD-1931/RDV and EIDD-1931/BRT combinations at the OACD and $C_{max}$ ratios (purple and pink markers, respectively) were also added to the infected Vero E6 cells and incubated for 72 h. The %Inhibition resulted from the mono- and combination therapies were measured via the luminescence signals of the cell viability in the ATP activity assay. The y-axis of each curve represent the %Inhibition for the each drug and combination. The $EC_{50}$ values for monotherapies are also summarized in the legends. Vertical dotted lines represent the 10% $C_{max}$ of EIDD-1931, and the horizontal dotted lines represent the $EC_{50}$ for monotherapies. No data points were excluded in dose-response curve analysis. Data points are presented as mean ± propagated SD (N = 3).

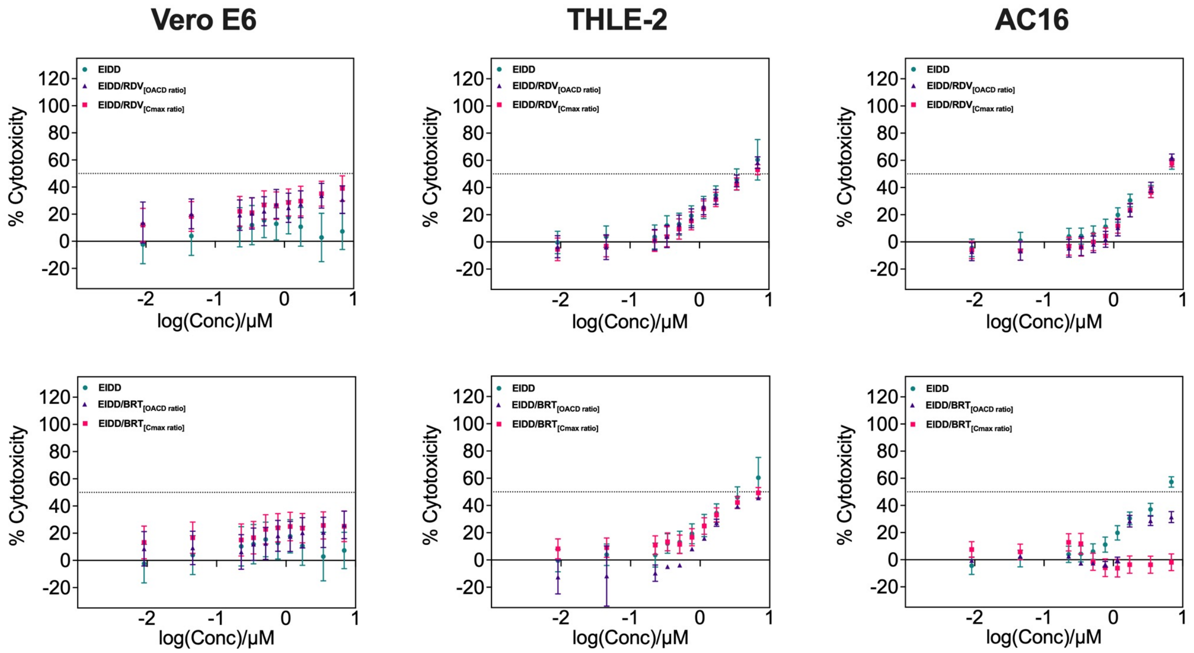


**Fig. S5. %Cytotoxicity data for EIDD-1931/RDV and EIDD-1931/BRT in Vero E6, THLE-2, and AC16 cell lines.** %Cytotoxicity for Vero E6 was determined after 72 h of treatment with EIDD-1931/RDV and EIDD-1931/BRT combinations at increasing concentrations using luminescence-based ATP activity assay. %Cytotoxicity data (mean ± propagated SD, N=3 to 4) for EIDD-1931/RDV and EIDD-1931/BRT at two different ratios: OACD ratio (Level 2/Level 2 ratio for EIDD-1931/RDV and Level 1/Level 2 ratio for EIDD-1931/BRT from the IDentif.AI experimental set; purple triangles) and $C_{max}$ ratio ($C_{max}$/$C_{max}$; pink squares) of the two drugs in the combination. Additionally, %Cytotoxicity was measured in THLE-2 human liver cell lines and AC16 human cardiomyocyte. EIDD-1931/RDV and EIDD-1931/BRT were added to the cells for 72 h before measuring the cell viability via luminescence-based ATP activity assay. Remdesivir (RDV), and baricitinib (BRT).

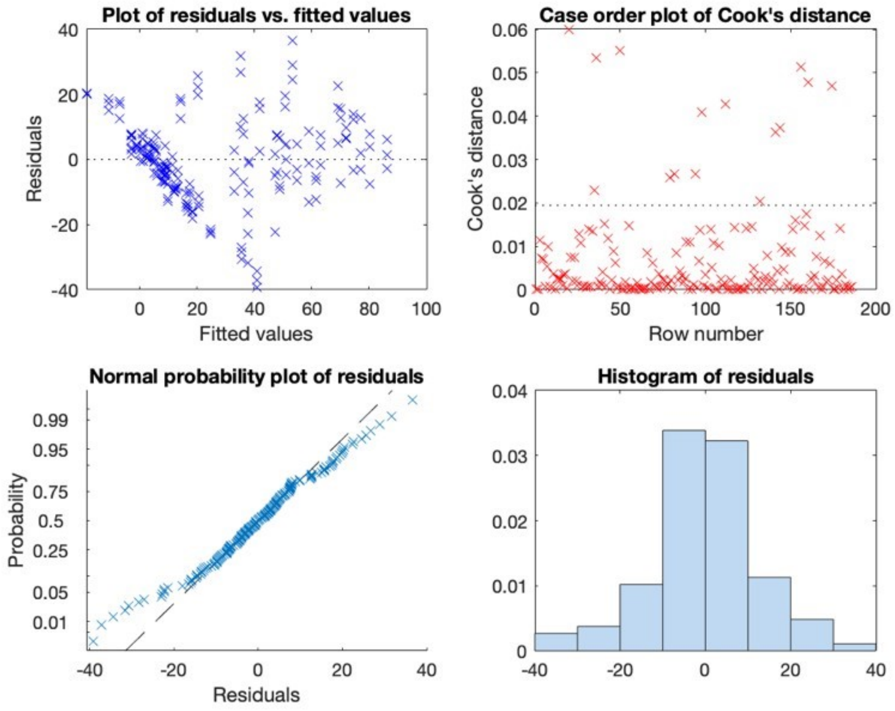


**Fig. S6. Outlier analysis for individual replicates in IDentif.AI %Inhibition analysis.** All experimental replicates (N = 3) of the %Inhibition of OACD-designed combinations and drug monotherapies were used in a quadratic stepwise regression analysis. Residuals represent the difference between the experimentally determined %Inhibition and the IDentif.AI-determined %Inhibition. The plot of residuals vs. fitted values examined the distributions of residuals and the quadratic model fit. Row number in the Cook's distance plot represents each OACD combination and monotherapy (triplicates) in order. The normal probability plot and the histogram of residuals were used to assess the normality of residual distribution. No data points were removed for the IDentif.AI analysis.

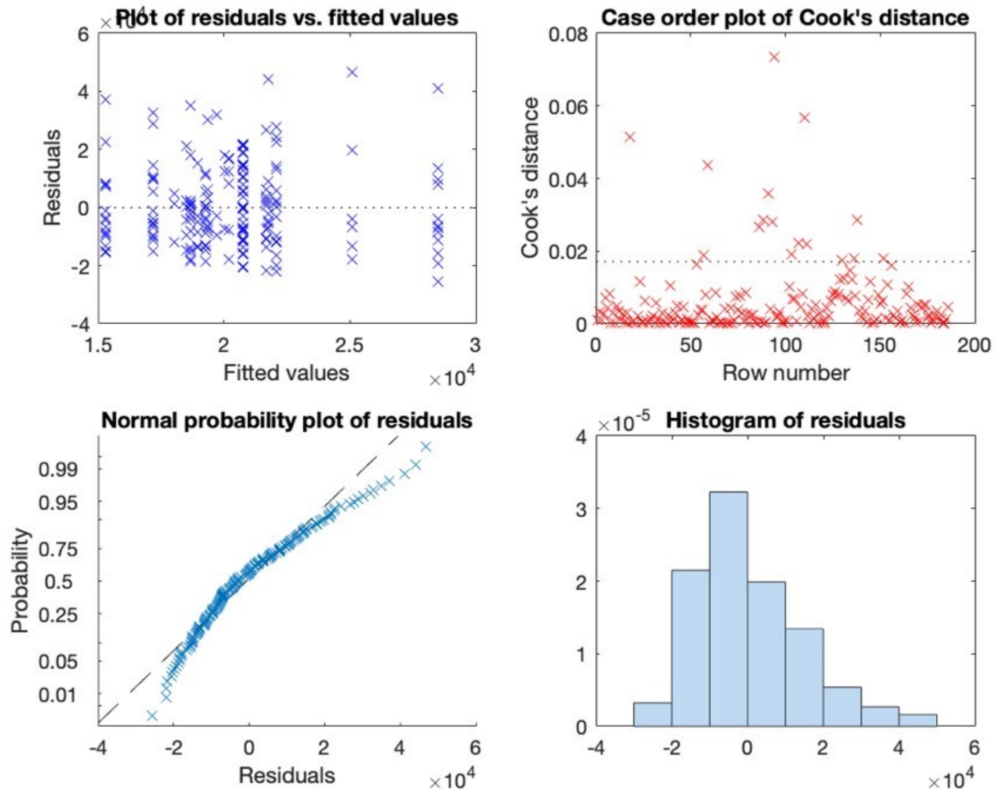


**Fig. S7. Outlier analysis for individual replicates in IDentif.AI Vero E6 %Cytotoxicity analysis.** All experimental replicates (N = 3) of the %Cytotoxicity of OACD-designed combinations and drug monotherapies were used in a quadratic stepwise regression analysis. Residuals represent the difference between the experimentally determined %Inhibition and the IDentif.AI-determined %Inhibition. The plot of residuals vs. fitted values examined the distributions of residuals and the quadratic model fit. Row number in the Cook's distance plot represents each OACD combination and monotherapy (triplicates) in order. The normal probability plot and the histogram of residuals were used to assess the normality of residual distribution. No data points were removed for the IDentif.AI analysis.

**Table S1. Resolution VI 6-drug OACD design.** 50 combinations for 6-drug library at three different concentration levels. Level 0 (input -1 in the OACD table) indicates the absence of a drug. Level 1 and Level 2 (inputs 0 and 1 in the OACD table, respectively) represent two clinically actionable drug concentrations. Remdesivir (RDV), ebselen (EBS), masitinib (MST), imatinib mesylate (IMT), and baricitinib (BRT).

| Combination | RDV | EBS | MST | IMT | BRT | EIDD-1931 |
|---|---|---|---|---|---|---|
| 1 | -1 | -1 | -1 | -1 | -1 | -1 |
| 2 | 1 | -1 | -1 | -1 | -1 | 1 |
| 3 | -1 | 1 | -1 | -1 | -1 | 1 |
| 4 | 1 | 1 | -1 | -1 | -1 | -1 |
| 5 | -1 | -1 | 1 | -1 | -1 | 1 |
| 6 | 1 | -1 | 1 | -1 | -1 | -1 |
| 7 | -1 | 1 | 1 | -1 | -1 | -1 |
| 8 | 1 | 1 | 1 | -1 | -1 | 1 |
| 9 | -1 | -1 | -1 | 1 | -1 | 1 |
| 10 | 1 | -1 | -1 | 1 | -1 | -1 |
| 11 | -1 | 1 | -1 | 1 | -1 | -1 |
| 12 | 1 | 1 | -1 | 1 | -1 | 1 |
| 13 | -1 | -1 | 1 | 1 | -1 | -1 |
| 14 | 1 | -1 | 1 | 1 | -1 | 1 |
| 15 | -1 | 1 | 1 | 1 | -1 | 1 |
| 16 | 1 | 1 | 1 | 1 | -1 | -1 |
| 17 | -1 | -1 | -1 | -1 | 1 | 1 |
| 18 | 1 | -1 | -1 | -1 | 1 | -1 |
| 19 | -1 | 1 | -1 | -1 | 1 | -1 |
| 20 | 1 | 1 | -1 | -1 | 1 | 1 |
| 21 | -1 | -1 | 1 | -1 | 1 | -1 |
| 22 | 1 | -1 | 1 | -1 | 1 | 1 |
| 23 | -1 | 1 | 1 | -1 | 1 | 1 |
| 24 | 1 | 1 | 1 | -1 | 1 | -1 |
| 25 | -1 | -1 | -1 | 1 | 1 | -1 |
| 26 | 1 | -1 | -1 | 1 | 1 | 1 |
| 27 | -1 | 1 | -1 | 1 | 1 | 1 |
| 28 | 1 | 1 | -1 | 1 | 1 | -1 |
| 29 | -1 | -1 | 1 | 1 | 1 | 1 |
| 30 | 1 | -1 | 1 | 1 | 1 | -1 |

| | | | | | | |
|---|---|---|---|---|---|---|
| 31 | -1 | 1 | 1 | 1 | 1 | -1 |
| 32 | 1 | 1 | 1 | 1 | 1 | 1 |
| 33 | -1 | -1 | -1 | -1 | -1 | -1 |
| 34 | -1 | 0 | 0 | 0 | 0 | 0 |
| 35 | -1 | 1 | 1 | 1 | 1 | 1 |
| 36 | 0 | -1 | -1 | 0 | 0 | 1 |
| 37 | 0 | 0 | 0 | 1 | 1 | -1 |
| 38 | 0 | 1 | 1 | -1 | -1 | 0 |
| 39 | 1 | -1 | 0 | -1 | 1 | 0 |
| 40 | 1 | 0 | 1 | 0 | -1 | 1 |
| 41 | 1 | 1 | -1 | 1 | 0 | -1 |
| 42 | -1 | -1 | 1 | 1 | 0 | 0 |
| 43 | -1 | 0 | -1 | -1 | 1 | 1 |
| 44 | -1 | 1 | 0 | 0 | -1 | -1 |
| 45 | 0 | -1 | 0 | 1 | -1 | 1 |
| 46 | 0 | 0 | 1 | -1 | 0 | -1 |
| 47 | 0 | 1 | -1 | 0 | 1 | 0 |
| 48 | 1 | -1 | 1 | 0 | 1 | -1 |
| 49 | 1 | 0 | -1 | 1 | -1 | 0 |
| 50 | 1 | 1 | 0 | -1 | 0 | 1 |

**Table S2. IDentif.AI estimated coefficients for %Inhibition analysis.** Remdesivir (RDV), ebselen (EBS), masitinib (MST), imatinib mesylate (IMT), and baricitinib (BRT). Statistical significance was determined using F-test. *p < 0.05, **p < 0.01 and, *** p < 0.001.

| | **Estimate** | **Statistical Significance** |
|---|---|---|
| Intercept | 21.703 | *** |
| RDV | 7.458 | *** |
| EBS | -4.917 | *** |
| MST | -0.043 | |
| IMT | 0.011 | |
| BRT | -4.930 | *** |
| EIDD-1931 | 22.654 | *** |
| RDV:MST | -3.417 | ** |
| RDV:IMT | -1.955 | |
| RDV:EIDD-1931 | 6.646 | *** |
| EBS:MST | -2.465 | * |
| EBS:BRT | -3.878 | *** |
| EBS:EIDD-1931 | -4.004 | *** |
| BRT:EIDD-1931 | -4.954 | *** |
| $MST^2$ | 6.035 | |
| $BRT^2$ | 19.688 | *** |
| $EIDD\text{-}1931^2$ | -16.002 | *** |
| Adj $R^2$ (IDentif.AI) | 0.794 | |

**Table S3. IDentif.AI estimated coefficients for Vero E6 %Cytotoxicity analysis.** Remdesivir (RDV), ebselen (EBS), masitinib (MST), imatinib mesylate (IMT), and baricitinib (BRT). Statistical significance was determined using F-test. *P < 0.05, **P < 0.01, and *** P < 0.001.

| | Estimate | Statistical Significance |
|---|---|---|
| Intercept | 20426.0 | *** |
| RDV | 1460.3 | |
| MST | 1076.5 | |
| IMT | 1321.9 | |
| RDV:MST | 2125.8 | |
| RDV:IMT | 2072.3 | |
| Adj $R^2$ (IDentif.AI) | 0.017 | |

### 3. References


Blasiak A, Lim JJ, Seah SGK, Kee T, Remus A, Chye DH, et al. IDentif.AI: Rapidly optimizing combination therapy design against severe Acute Respiratory Syndrome Coronavirus 2 (SARS-CoV-2) with digital drug development. Bioeng Transl Med 2021;6:e10196. https://doi.org/10.1002/btm2.10196.

EMA 2005. Norvir: European Public Assessment Preport - Scientific Discussion. https://www.ema.europa.eu/en/documents/scientific-discussion/norvir-epar-scientific-discussion_en.pdf

EMA 2011. Kaletra: Summary of Product Characteristics. https://www.ema.europa.eu/en/documents/product-information/kaletra-epar-product-information_en.pdf

EMA 2017. Masipro: Assessment Report. https://www.ema.europa.eu/en/documents/assessment-report/masipro-epar-refusal-public-assessment-report_en.pdf

Farrance I, Frenkel R. Uncertainty of Measurement: A Review of the Rules for Calculating Uncertainty Components through Functional Relationships. Clin Biochem Rev 2012;33:49–75.

FDA 2002. Camptosar (irinotecan hydrochloride injection). https://www.accessdata.fda.gov/drugsatfda_docs/label/2002/20571s16lbl.pdf

FDA 2011. INCIVEK.

https://www.accessdata.fda.gov/drugsatfda_docs/label/2011/201917lbl.pdf

FDA 2018a. Olumiant: Pharmacology/Toxicology Review 2018a. https://www.accessdata.fda.gov/drugsatfda_docs/nda/2018/207924Orig1s000PharmR.pdf

FDA 2018b.Selinexor: Multi-Discipline Review https://www.accessdata.fda.gov/drugsatfda_docs/nda/2019/212306Orig1s000MultidisciplineR.pdf

FDA 2020. Veklury (remdesdivir) EUA Fact Sheet for Healthcare Providers. https://www.fda.gov/media/137566/download

KEGG 2019. Fusan (Nafamostat Mesilate). https://www.kegg.jp/medicus-bin/japic_med?japic_code=00048710

Kil J, Lobarinas E, Spankovich C, Griffiths SK, Antonelli PJ, Lynch ED, et al. Safety and efficacy of ebselen for the prevention of noise-induced hearing loss: a randomised, double-blind, placebo-controlled, phase 2 trial. Lancet 2017;390:969–79. https://doi.org/10.1016/S0140-6736(17)31791-9.

Nikolova Z, Peng B, Hubert M, Seiberling M, Keller U, Ho YY, et al. Bioequivalence, safety, and tolerability of imatinib tablets compared with capsules. Cancer Chemother Pharmacol 2004;53:433–8. https://doi.org/10.1007/s00280-003-0756-z.

Painter WP, Holman W, Bush JA, Almazedi F, Malik H, Eraut NCJE, et al. Human safety, tolerability, and pharmacokinetics of molnupiravir, a novel broad-spectrum oral antiviral agent with activity against SARS-CoV-2. Antimicrob Agents Chemother 2021;65. https://doi.org/10.1128/AAC.02428-20.

Xu H, Jaynes J, Ding X. Combining two-level and three-level orthogonal arrays for factor screening and response surface exploration. Stat Sin 2014;24:269–89. https://doi.org/10.5705/ss.2012.210.

Young BE, Ong SWX, Kalimuddin S, Low JG, Tan SY, Loh J, et al. Epidemiologic Features and Clinical Course of Patients Infected With SARS-CoV-2 in Singapore. JAMA 2020;323:1488–94. https://doi.org/10.1001/jama.2020.3204.
